# Derivation of a prognostic model for critically ill children in locations with limited resources

**DOI:** 10.1101/2023.05.19.23290233

**Authors:** Arjun Chandna, Suy Keang, Meas Vorlark, Bran Sambou, Chhay Chhingsrean, Heav Sina, Pav Vichet, Kaajal Patel, Eang Habsreng, Arthur Riedel, Lazaro Mwandigha, Constantinos Koshiaris, Rafael Perera-Salazar, Paul Turner, Ngoun Chanpheaktra, Claudia Turner

**Affiliations:** Cambodia Oxford Medical Research Unit, Angkor Hospital for Children, Siem Reap, Cambodia; Centre for Tropical Medicine & Global Health, University of Oxford, Oxford, United Kingdom; Department of Intensive Care Medicine, Angkor Hospital for Children, Siem Reap, Cambodia; Department of Global Child Health, Angkor Hospital for Children, Siem Reap, Cambodia; Department of Primary Care Health Sciences, University of Oxford, Oxford, United Kingdom; Angkor Hospital for Children, Siem Reap, Cambodia

**Keywords:** Paediatrics, critical care, intensive care, prediction model, risk stratification, resource-limited

## Abstract

**Background:** Capacity and demand for paediatric critical care are growing in many resource-constrained contexts. However, tools to support resource stewardship and promote sustainability of critical care services are lacking.

**Methods:** This study assessed the ability of nine severity scores to risk stratify children admitted to a paediatric intensive care unit (PICU) in Siem Reap, northern Cambodia. It then developed a bespoke clinical prediction model to enable risk stratification in resource-constrained PICU contexts. The primary outcome was death during PICU admission.

**Results:** 1,550 consecutive PICU admissions were included, of which 97 (6.3%) died. Most existing severity scores achieved comparable discrimination (area under the receiver operating characteristic curves [AUCs] 0.71-0.76) but only three scores demonstrated moderate diagnostic utility for triaging admissions into high- and low-risk groups (positive likelihood ratios 2.65-2.97 and negative likelihood ratios 0.40-0.46). The newly derived model outperformed all existing severity scores (AUC 0.84, 95% CI 0.80-0.88; p < 0.001). Using one particular threshold, the model classified 13.0% of admissions as high-risk, amongst which probability of mortality was almost ten-fold greater than admissions triaged as low-risk (PLR 5.75; 95% CI 4.57-7.23 and NLR 0.47; 95% CI 0.37-0.59). Decision curve analyses indicated that the model would be superior to all existing severity scores and could provide utility across the range of clinically plausible decision thresholds.

**Conclusions:** Existing paediatric severity scores have limited potential as risk stratification tools in resource-constrained PICUs. If validated, the prediction model developed herein would provide a readily implementable mechanism to support triage of critically ill children on admission to PICU and could be tailored to suit a variety of contexts where resource prioritisation is important.

## Introduction

Historically, paediatric critical care has often been perceived as too complex, expensive, or unethical to provide in settings where resources are scarce.^1^ These presumptions are countered by data which suggest that simple, low-cost interventions can result in substantial improvements in health outcomes and the premise of comprehensive healthcare as a universal human right.^2–5^ Consequently, demand and capacities for paediatric critical care are growing in many resource-limited settings.^6, 7^

Notwithstanding these welcome developments, need for critical care often outstrips supply.^8^ Evidence-based approaches to support resource stewardship are essential to promote equitable and sustainable critical care services. This is especially true in rural regions of many low- and middle-income countries (LMICs) where maldistribution of healthcare professionals and resources results in considerable disparities in access to paediatric critical care.^1, 3, 4, 9–12^

Risk stratification tools can help target scarce resources optimally. However, tools developed for use in paediatric intensive care units (PICUs) in high-income settings are time-consuming to compute and require diagnostic tests not routinely available in resource-constrained regions of LMICs.^13, 14^ Furthermore, prognosis is influenced by the level of care available and underlying host susceptibility states, and hence adapted tools are required to support context-specific clinical decision making.^9, 15, 16^ Consequently, there have been calls both to validate existing severity scores in resource-constrained PICUs and to develop new risk stratification tools appropriate for use in these settings.^9, 17^ Unfortunately, most studies from LMIC PICUs to date have been limited to urban centres, hampered by small sample sizes, and used methods incompatible with development of robust clinical severity scores or prediction models.^18–22^

Using data from children admitted to the PICU at the Angkor Hospital for Children (AHC) in Siem Reap, Cambodia, this study reports the external validation of nine existing paediatric severity scores in a resource-limited PICU setting. Secondly, it presents the development of a bespoke clinical prediction model, derived specifically to support risk stratification in resource-constrained PICU contexts.

## Methods

This retrospective cohort study screened consecutive admissions to the PICU at AHC between 01/01/2018 and 01/01/2020. All non-elective admissions of children aged > 28 days and ≤ 16 years were included. The study was approved by the AHC Research Committee (AHC 0656/20),

Cambodian National Ethics Committee for Health Research (NECHR 257), and the Oxford Tropical Research and Ethics Committee (OxTREC 565-20), and is reported in accordance with the Transparent Reporting of a multivariable prediction model for Individual Prognosis Or Diagnosis (TRIPOD) guidelines (Appendix 1).^22^

### Study setting

AHC is a non-governmental paediatric healthcare organisation with a nationwide catchment area providing comprehensive primary-to-tertiary care. The hospital, located in Siem Reap, northern Cambodia, has 89 inpatient beds situated on two medical wards, a surgical ward, a special care baby unit, and neonatal and paediatric intensive care units. The 14-bedded PICU has approximately 1,000 annual admissions and is staffed by a team of 30 nurses, four senior doctors, five doctors in training, and receives medical and nursing students from Cambodia’s three main medical schools. The unit provides the only critical care service for children in the north of the country and is the only PICU located outside of the capital city, Phnom Penh. Clinical staff have completed or are undertaking training in paediatric intensive care medicine. The unit (Level II or Community PICU)^11, 23^ provides mechanical and non-invasive ventilation (oxygen cylinders are delivered fortnightly), inotropic therapy, peritoneal dialysis, and specialist nursing care (minimum 1:3 nurse-patient ratio) for critically ill children at AHC and those transferred from other health facilities. A backup generator ensures continuity of electrical supply during infrequent power outages.

### Data collection

PICU admissions were identified from the electronic Hospital Information System (HIS) and cross-checked against the admission logbook located on the unit. Clinical records were retrieved and data extracted onto structured case report forms (CRFs) by a team of research nurses/assistants who had been trained by a Principal Investigator (AC). Data extraction occurred between 27/11/2020 and 14/12/2021. It was not possible to blind the research team to outcome status during data extraction. The hospital admission and PICU vital sign proforma (Appendix 2) helped standardise data extraction and all variables were prospectively defined in a data dictionary to ensure consistency of interpretation across the research team. Each CRF was reviewed by a study doctor (AC or SK) in consultation with the clinical records, with particular focus on explanatory and outcome variables. Data were entered into an electronic study database and 10% of CRFs underwent review by the study Data Manager (VP) to ensure a data entry error rate of < 0.5%. Data profiling was conducted to identify missing and implausible values.

### Shortlisting of existing severity scores

The results of two recent systematic reviews were supplemented by searching PubMed using synonyms of “paediatric” AND “severity score OR prediction model”.^24, 25^ Forty-nine scores or models were longlisted and assessed for suitability for external validation (Appendix 3): 15 were excluded as they required diagnostic tests unavailable in resource-limited PICU settings (for example, arterial blood gases, creatinine, or serum electrolyte estimations) and 14 were excluded as they contained variables not relevant to the intended-use context and/or population (for example, arrival via emergency medical services, presence of an indwelling central venous catheter). A further five were excluded as the information required to calculate the score was not provided in the original manuscript, and another eight were excluded as they contained variables not available in the AHC clinical records and no suitable proxy variables could be identified. Ultimately, nine severity scores were shortlisted for external validation (Table 1). Neither the setting, population, nor outcome used for the derivation were prerequisites for selection of a score for external validation.

**Table 1.** Severity scores selected for external validation. Scores were selected for external validation irrespective of the setting, population, and outcome used for the original derivation study. The only prerequisites were that the score had to be calculable with the available data (with the exception that systolic blood pressure could be dropped if CRT was included), relevant to the study population, and feasible for implementation in a resource-limited PICU context. AVPU = Alert Voice Pain Unresponsive scale; CRT = capillary refill time; ED = emergency department; FEAST-PET = Fluid Expansion as Supportive Therapy-Pediatric Emergency Triage; GCS = Glasgow Coma Scale; LqSOFA = Liverpool quick Sequential Organ Failure Assessment; PAWS = Paediatric Advanced Warning Score; PEWS = Pediatric Early Warning System; PEWS-RL = PEWS-Resource Limited; PICU = paediatric intensive care unit; qPELOD-2 = quick Pediatric Logistic Organ Dysfunction-2; SIRS = systemic inflammatory response syndrome; SpO_2_ = oxygen saturation.

| Score | Range | Predictors | Adjustments | Original setting, population, and outcome |
| --- | --- | --- | --- | --- |
| **FEAST-PET** | 0-10 | Heart rate, temperature, CRT, pulse character, work of breathing, lung crepitations, mental status, pallor | Cut-off for CRT increased to $\geq 3$ seconds; deep breathing omitted from work of breathing; age-adjusted WHO criteria for severe anaemia used as a proxy for pallor;<sup>55</sup> mental status dichotomised and assessed using AVPU or GCS, reducing the maximum possible score to 9 | Score to predict 48-hour mortality on admission to secondary and tertiary-care hospitals in East Africa in children with severe febrile illness<sup>41</sup> |
| **LqSOFA** | 0-4 | Respiratory rate, heart rate, CRT, mental status | Mental status assessed using AVPU or GCS | Score to predict PICU admission or death in febrile children presenting to ED in the United Kingdom<sup>43</sup> |
| **PAWS** | 0-21 | Respiratory rate, heart rate, temperature, CRT, mental status, SpO<sub>2</sub>, work of breathing | Mental status assessed using AVPU or GCS; CRT, mental status, and work of breathing dichotomised, reducing the maximum possible score to 17 | Score to predict need for PICU admission in children presenting to ED in the United Kingdom<sup>45</sup> |
| **PEWS** | 0-26 | Respiratory rate, heart rate, systolic blood pressure, CRT, SpO<sub>2</sub>, supplemental oxygen, work of breathing | Systolic blood pressure omitted; work of breathing dichotomised, reducing the maximum possible score to 20 | Score to predict need for PICU admission in children hospitalised on a general paediatric ward in a tertiary-care hospital in Canada<sup>46</sup> |
| **PEWS-IRISH** | 0-21 | Respiratory rate, heart rate, systolic blood pressure, CRT, mental status, SpO<sub>2</sub>, supplemental oxygen, work of breathing | Systolic blood pressure omitted; work of breathing and mental status dichotomised, reducing the maximum possible score to 15 | Adaptation of the PEWS score by the Irish National Clinical Effectiveness Committee<sup>56</sup> |
| **PEWS-RL** | 0-6 | Respiratory rate, heart rate, temperature, mental status, supplemental oxygen, work of breathing |  | Score to predict clinical deterioration in children hospitalised on a general paediatric ward in a tertiary-care hospital in Rwanda<sup>47</sup> |
| **qPELOD-2** | 0-3 | Heart rate, systolic blood pressure, mental status | CRT used as a proxy for systolic blood pressure; mental status assessed using AVPU or GCS | Score to predict mortality in children with suspected infection on admission to nine European PICUs<sup>38</sup> |
| **qSOFA** | 0-3 | Respiratory rate, systolic blood pressure, mental status | CRT used as a proxy for systolic blood pressure; mental status assessed using AVPU or GCS | Adult sepsis score to predict mortality adapted for children with suspected infection on admission to PICUs in Australia and New Zealand<sup>38</sup> |
| **SIRS** | 0-4 | Respiratory rate, heart rate, temperature, white cell count |  | Expert consensus definition for the diagnosis of paediatric sepsis<sup>57</sup> |

### Candidate predictors

Baseline variables at the time of PICU admission were extracted from the clinical records. For admissions occurring from the AHC Emergency Room (ER) the first set of vital signs was abstracted. For inter- and intra-hospital transfers the vital signs recorded at the time the decision to transfer was made were abstracted. If weight and height were not recorded at the time of PICU admission the closest values during the same hospital stay were used. Laboratory parameters measured within 24 hours of PICU admission were considered available on admission. A sensitivity analysis restricting this period to between two hours prior and up to four hours after admission was performed (Appendix 4).^26^

For derivation of the new model, candidate predictors were selected a priori based on existing literature, expert knowledge, feasibility for implementation, and availability of data in the clinical records. Variables were divided into five ‘domains’: background, illness journey, cardiovascular, respiratory, and neurological. Candidate predictors were selected across all domains to ensure holistic assessment of critical illness and inclusion of important contextual determinants of outcome often neglected by clinical risk scores developed in high-income settings. The 11 selected predictors were age, comorbidity status, weight-for-age z-score, estimated travel time to hospital, route of admission to PICU, heart rate, central capillary refill time (CRT), respiratory rate, oxygen saturation (SpO_2_), receipt of supplemental oxygen, and mental status.

### Outcomes

The primary outcome was death during PICU admission. Participants who were discharged from PICU to die at home were classified as meeting the primary outcome. A sensitivity analysis was conducted excluding these participants as well as those whose death was judged by either of the study doctors (AC or SK) to have been related to a separate illness acquired during the PICU stay (Appendix 4).

The secondary outcome was death in the 12 months following a PICU discharge. Caretakers of participants for whom post-discharge outcomes could not be determined from the clinical records or HIS were telephoned to ascertain vital status 12 months after PICU discharge.

### Sample size

Routinely collected data from the hospital indicated that approximately 100 deaths on the PICU were expected over two calendar years (mortality rate of ∼5%), which would ensure sufficient outcome events for external validation of the existing severity scores.^27^ At this prevalence, and assuming a conservative Nagelkerke R^2^ of 0.15, up to 10 candidate predictors (events per parameter [EPP] = 9.7) could be used to build the new prediction model (R package: *pmsampsize*).^28, 29^ In order to allow for inclusion of interaction terms between age and heart rate and age and respiratory rate penalisation was used to shrink regression coefficients and permit inclusion of up to 13 parameters whilst still minimising the risk of overfitting.

### Missing data

Missing data were summarised for the existing scores and for each candidate predictor in the new model (Appendix 5; R package: *naniar*).^30^ For the existing scores, missingness ranged from 5.4% (qPELOD-2) to 15.6% (FEAST-PET). Amongst the 11 candidate predictors for the new model missingness ranged from 1.1% for heart rate and prolonged CRT to 6.9% for respiratory rate, whilst four predictors had no missing data. Given the relatively low proportion of missing data, single (median) imputation grouped by outcome status was proposed to address missingness. Sensitivity analyses comparing this to a full-case approach, as well as best- and worst-case imputation, produced similar results (Appendix 5), confirming that single (median) imputation was appropriate for the primary analysis.

### Statistical methods

Discrimination and calibration of each existing score was assessed by quantifying the area under the receiver operating characteristic curve (AUC; R package: *pROC*)^31^ and plotting the proportion of admissions that met the primary outcome at each level of a score. Positive and negative likelihood ratios (PLRs and NLRs) were reported at each of the scores’ cut-offs to quantify the change in pre-test probability that a PICU admission would result in death. As a rule-of-thumb, a PLR > 10 or NLR < 0.1 is often deemed conclusive, a PLR between 5-10 or NLR between 0.1-0.2 considered substantial, a PLR between 2-5 or NLR between 0.2-0.5 regarded as small but important, and a PLR between 1-2 or NLR between 0.5-1 likely clinically insignificant.^32^

Prior to model building the relationship between continuous predictors and PICU survival status was examined using a loess smoothing approach to determine if transformations were required. Age-specific relationships for heart rate and respiratory rate (< 12 months; 12-59 months; 5-12 years; > 12 years) were explored to account for known changes in these parameters associated with physiological maturation. Stratum-specific odds ratios (ORs) and likelihood ratio tests (LRTs) were used to identify important interactions between age and each of heart rate and respiratory rate, as well as between SpO_2_ and receipt of supplemental oxygen. Penalised (ridge) logistic regression was used to derive the model and adjust for optimism (R package: *ridge*).^33^ All predictors were prespecified and no predictor selection was performed during model development.

Discrimination (AUC), calibration (calibration intercept, slope, and plots), and classification indices at clinically-relevant decision thresholds (R package: *reportROC*)^34^ were reported to summarise model performance. Recognising that the relative value of a true positive (TP; admission correctly identified as at high-risk of death) and false positive (FP; admission incorrectly identified as at high-risk of death) will be context-dependent (for example, depending on the human and material capacities of a high-acuity area that at-risk admissions might be triaged to), the clinical utility of the new model was compared to the best-performing existing scores using decision curves to visualise their net-benefits over a range of clinically-plausible decision thresholds (R package: dcurves).^35, 36^

All analyses were done in R, version 4.2.2.^37^

## Results

### Study population

Between 01/01/2018 and 01/01/2020 there were 2,066 admissions to the hospital’s PICU, of which case notes were located for 2,021 (97.8%; 2,021/2,066). In total, 1,550 non-elective admissions were eligible for inclusion in the study (eligibility rate 76.7%; 1,550/2,021; Appendix 6). There were 1,366 unique children in the cohort, with 91.1% (1,245/1,366) admitted to the PICU only once during the study period. Median age at PICU admission was 14.0 months (interquartile range [IQR] 4.0-73.0 months) and 59.8% of admissions (927/1,550) were for male children (Table 2). Nearly one in five admissions had a WAZ < -3 (271/1,550; 17.5%) with a similar proportion having a WAZ between -3 and -2 (261/1,550; 16.8%).

**Table 2.**
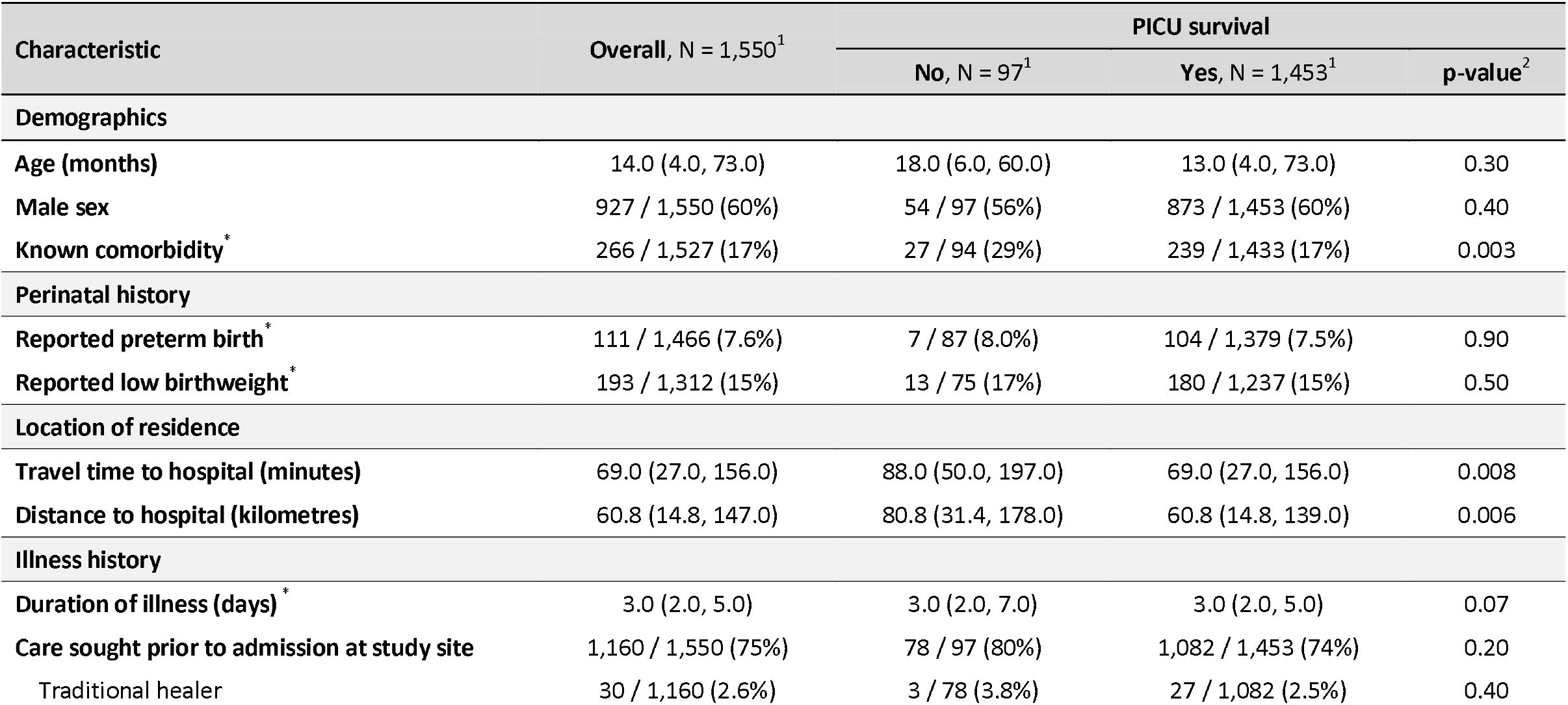
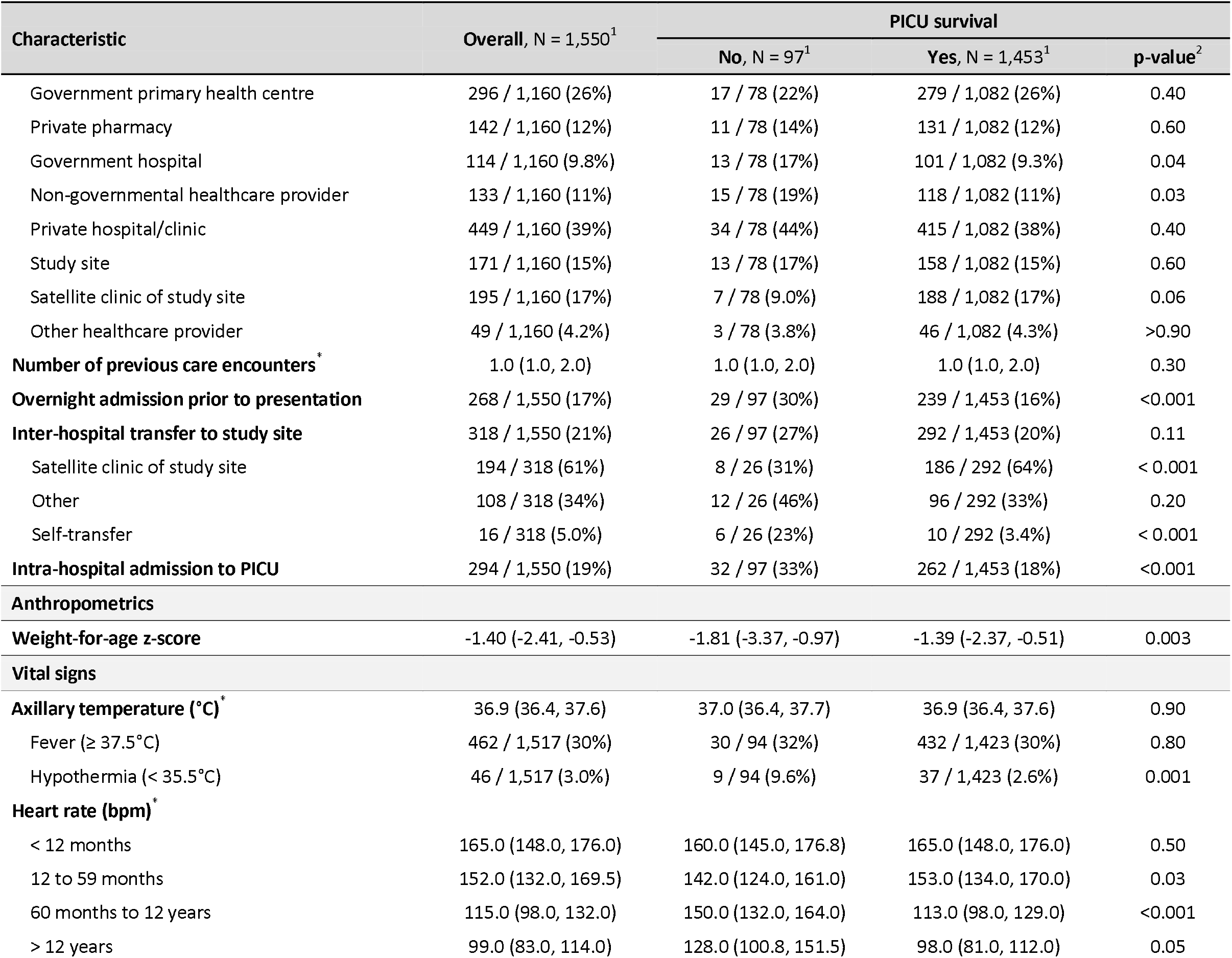
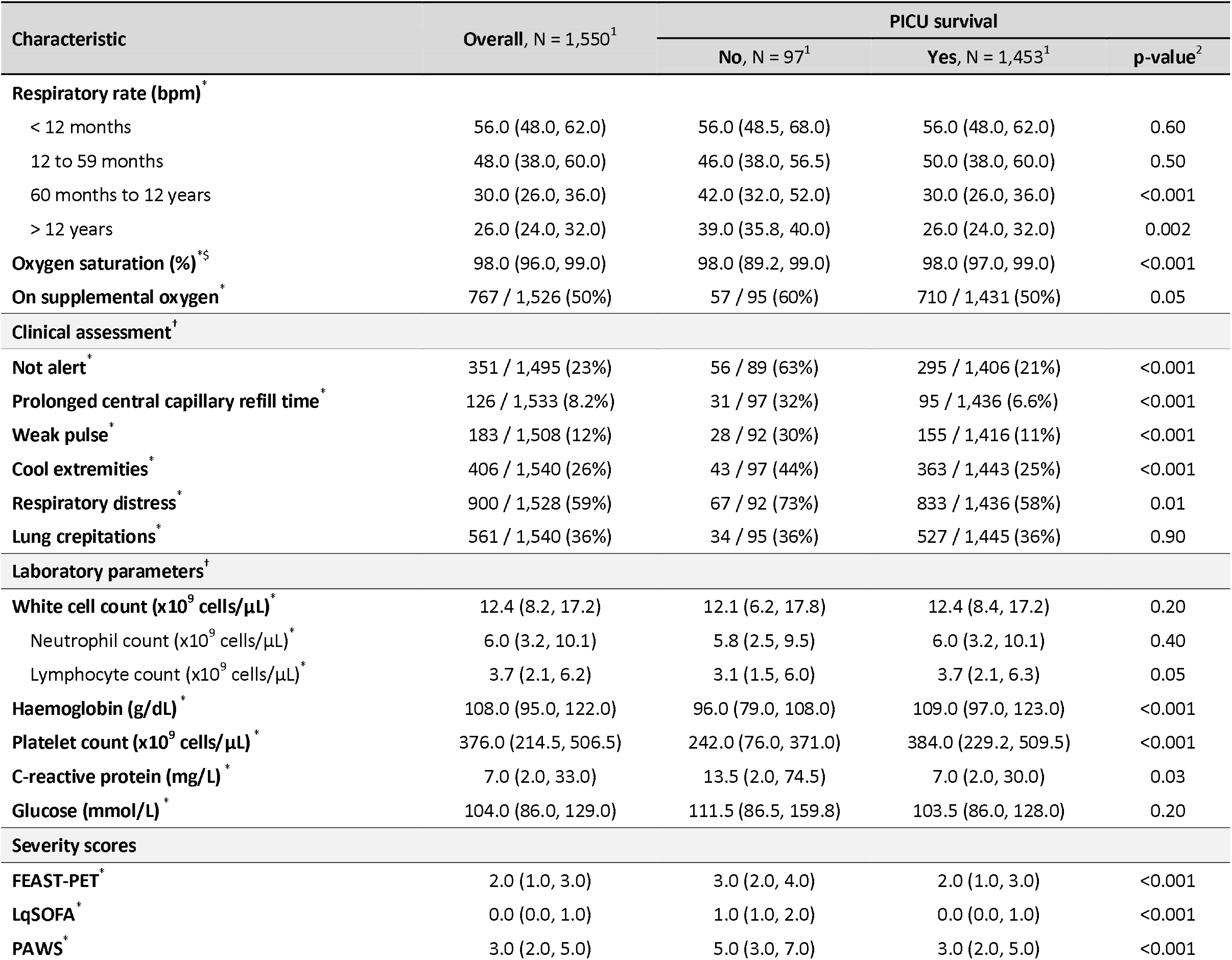
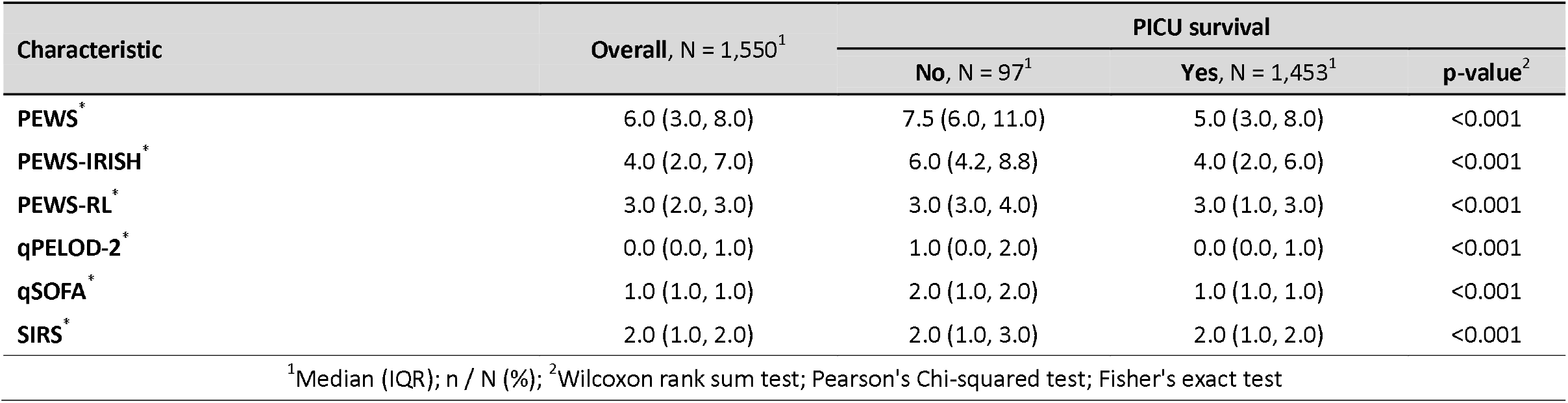
Baseline characteristics. Baseline demographic, background, illness history, anthropometric, clinical, and laboratory characteristics of the cohort, stratified by primary outcome status. *Missing data: comorbidity = 23; preterm birth = 84; low birthweight = 238; illness duration = 2; number of previous care encounters = 391; axillary temperature = 33; heart rate = 17; respiratory rate = 107; oxygen saturation = 26; supplemental oxygen = 24; mental status = 55; CRT = 17; pulse character = 42; cool extremities = 10; respiratory distress = 22; lung crackles = 10; white cell count = 117; neutrophil count = 117; lymphocyte count = 118; haemoglobin = 114; platelet count = 115; C-reactive protein = 411; glucose = 386; LqSOFA = 177; qSOFA = 170; qPELOD-2 = 83; SIRS = 235; PEWS = 175; PEWS-RL = 206; PEWS-IRISH = 220; PAWS = 209; FEAST-PET = 242. ^$^Baseline SpO amongst those not receiving supplemental oxygen at the time of PICU admission confirmed a similar relationship (96.5% vs. 98.0%; p < 0.001; n = 798). ^†^Not alert = GCS < 15 or AVPU < A; prolonged CRT defined as > 2 seconds; respiratory distress = chest indrawing, tracheal tug, or nasal flaring; laboratory parameters included if measured within 24 hours of PICU admission. AVPU = Alert Voice Pain Unresponsiveness scale; Bpm = beats/breaths per minute; CRT = capillary refill time; GCS = Glasgow Coma Scale; PICU = paediatric intensive care unit.

### Illness journeys

Admissions originated from 23 of Cambodia’s 25 provinces (Appendix 7) and median travel time from a child’s residence to the hospital was 69 minutes (IQR 27-156 minutes). Children had been sick for a median of three days (IQR 2-5 days) prior to admission to the study site, with the majority (1,160/1,550; 74.8%) seeking care from at least one other healthcare provider prior to presentation. Approximately one in five children (268/1,550; 17.3%) had been admitted overnight at another healthcare facility. A similar proportion were inter-hospital transfers (318/1,550; 20.5%), the majority referred from the hospital’s satellite clinic (194/318; 61.0%) located approximately 45 minutes from the main site.

Over two-thirds of PICU admissions originated from the hospital’s ER (1,067/1,550; 68.8%), whilst 294 (294/1,550; 19.0%) were intra-hospital transfers from one of the hospital’s three acute wards. Most children were admitted with febrile illnesses (1,128/1,540; 79.1%). Common reasons for PICU admission included respiratory distress (989/1,550; 63.8%), circulatory instability (545/1,550; 35.2%), and impaired consciousness (352/1,550; 22.7%).

### Baseline characteristics

Children with known comorbidities, as well as those sick for longer, admitted elsewhere prior to presentation, residing further from the hospital, and intra-hospital transfers were all less likely to survive their PICU admission (p < 0.001 to 0.07). Presentations with hypothermia, lower SpO_2_, impaired consciousness, and signs of cardiovascular (prolonged CRT, weak pulses, or cool extremities) or respiratory compromise were all more likely to meet the primary outcome (p < 0.001 to 0.01). Baseline tachycardia and tachypnoea were associated with death during PICU stay for children aged five years and older (p < 0.001 to 0.05) but this association was not observed for younger children. Statistically significant differences between admissions that did and did not meet the primary outcome were observed for a number of laboratory parameters, however only for haemoglobin was this association compatible with a potentially clinically important difference (96.0 vs. 109.0 g/dL; p < 0.001). All nine clinical severity scores were higher in admissions that resulted in death (p < 0.001).

### Clinical outcomes

Vital organ support was provided to 41.7% (647/1,550) of PICU admissions: 516 /1,550 (33.3%) were non-invasively ventilated, 354/1,550 (22.8%) mechanically ventilated, 98/1,550 (6.3%) received inotropic therapy, and 4/1,550 (0.3%) received peritoneal dialysis. Median length of stay on the unit was two days (IQR 1-4 days). The most frequent discharge diagnoses included pneumonia (501/1,550; 32.3%), bronchiolitis (319/1,550; 20.6%), and dengue (221/1,550; 14.3%).

The PICU mortality rate was 6.3% (97/1,550), with 85 children dying during their PICU stay and a further 12 discharged to die at home. The most common causes of death were pneumonia (52/97; 53.6%), undifferentiated sepsis (15/97; 15.5%), and cardiac failure (9/97; 9.3%), although 28 children (28/97; 28.9%) had more than one cause of death implicated (Appendix 8). Median time to death was four days (IQR 1-7 days; Appendix 9a).

Of the 1,453 admissions surviving to leave PICU, vital status 12 months post-PICU discharge could be ascertained for 782 (53.8%; 782/1,453). Of these, 33/782 admissions (4.2%) had resulted in death (25/672 [3.7%] individual children), a median of one month (IQR 1-4 months) post-discharge (Appendix 9b).

### External validation of existing severity scores

All scores achieved comparable discrimination (Figure 1a; AUCs 0.71-0.76) with the exception of SIRS, for which discrimination was poorer (AUC 0.59; 95% CI 0.53-0.65). Across all scores, admissions with higher baseline scores were more likely to progress to meet the primary outcome, although this association was less pronounced for PEWS-RL and SIRS (Appendix 10). For scores with multiple levels (PAWS, PEWS, and PEWS-IRISH) the increase in the proportion of admissions meeting the primary outcome across lower levels was modest, indicating redundancy in these more granular scoring systems.

**Figure 1.**
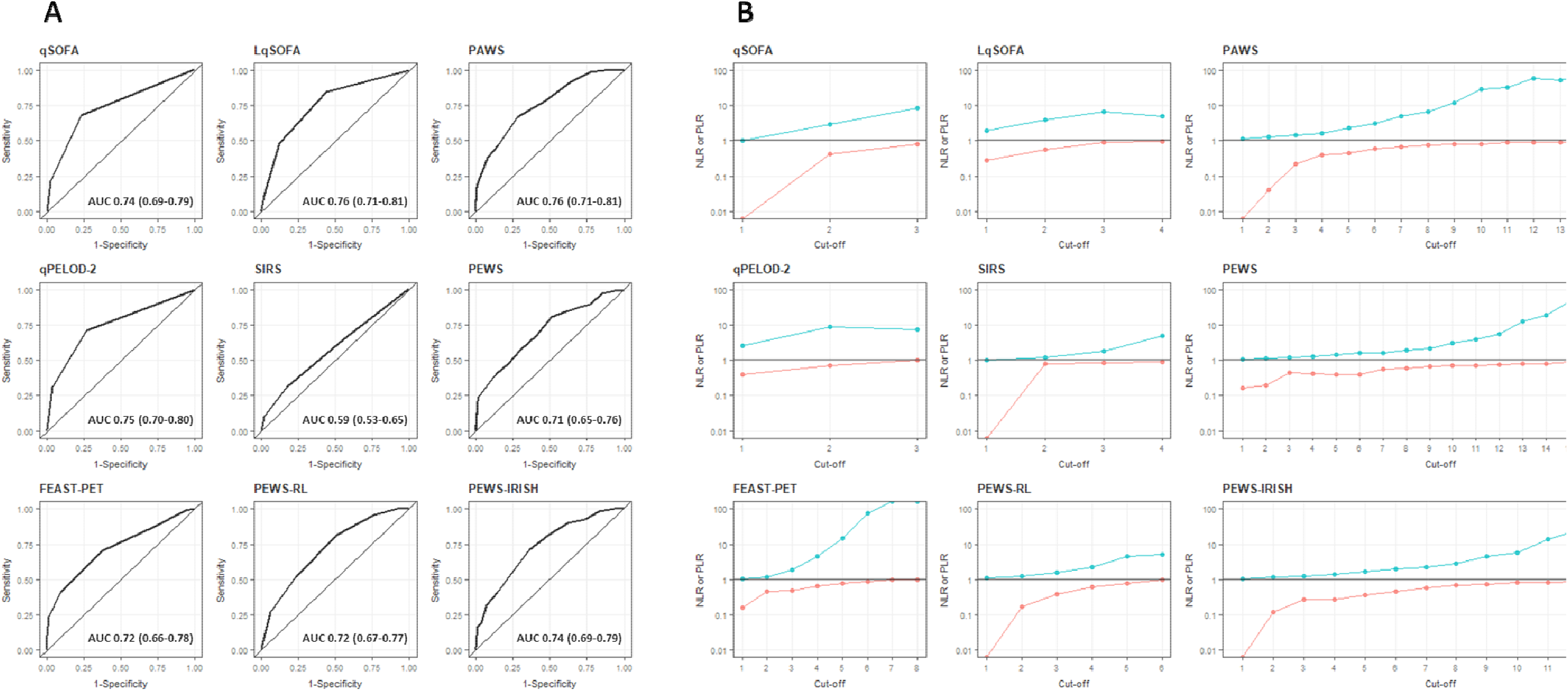
Performance of the nine existing severity scores. <u>Panel A</u>: discrimination of the severity scores. Perfect discrimination is indicated by an AUC of 1.0. <u>Panel B</u>: negative (red line) and positive (blue line) likelihood ratios of the severity scores at different cut-offs, illustrated on a log_10_ scale. As a rule-of-thumb, a greater than 10-fold change (PLR > 10 or NLR < 0.1) is often deemed conclusive, a 5-to-10-fold change (PLR 5-10 or NLR 0.1-0.2) substantial, a 2-to-5-fold change (PLR 2-5 or NLR 0.2-0.5) small but important, and a less than two-fold change (PLR 1-2 or NLR 0.5-1) likely clinically insignificant.^32^ AUC = area under the receiver operating characteristic curve; NLR = negative likelihood ratio; PLR = positive likelihood ratio.

At a cut-off of ≥ 1, the qPELOD-2 score demonstrated a sensitivity of 0.71 (95% CI 0.62-0.80) and specificity of 0.73 (95% CI 0.71-0.75). No other score achieved a sensitivity and specificity of > 0.70 at any cut-off (Appendix 11). Three scores demonstrated potential for stratifying PICU admissions into low- and high-risk groups (Figure 1b), achieving small but important changes in pre-test probability using a single cut-off: qPELOD-2 at a cut-off of ≥ 1 (PLR 2.65; 95% CI 2.28-3.09 and NLR 0.40; 95% CI 0.29-0.54), qSOFA at a cut-off of ≥ 2 (PLR 2.97; 95% CI 2.52-3.50 and NLR 0.42; 95% CI 0.31-0.56), and PAWS at a cut-off of ≥ 5 (PLR 2.40; 95% CI 2.04-2.82 and NLR 0.46; 95% CI 0.34-0.61).

### Derivation of a new prediction model for resource-constrained PICU contexts

Assessment of the relationship between continuous candidate predictors and the primary outcome did not identify serious violations of linearity (Appendix 12). Age-dependent relationships between the primary outcome and heart rate and respiratory rate were evident and this was confirmed via examination of stratum-specific ORs and LRTs for interaction (p < 0.001). Examination of stratum-specific ORs and an LRT for interaction between SpO_2_ and supplemental oxygen status did not indicate evidence of interaction (p = 0.92) and thus only the main effects for these parameters were included in the model. The full model, including the formulae to calculate the probability that a PICU admission will result in death, is presented (Table 3).

**Table 3.** Final clinical prediction model to estimate the probability that a PICU admission will end in death. Predictors spanning the five clinical domains are presented along with their regression coefficients and the formulae required to calculate the probability that a PICU admission will end in death.* Assessed by a Principal Investigator (CT) blinded to outcome status using the following adapted working definition: *any previous health condition known to be present at PICU admission severe enough to require specialty paediatric care and probably a period of hospitalisation over 12 months*.^58 #^ Calculated (R package: *zscorer*)^59^ using WHO (children < 10 years)^60, 61^ and US CDC (children ≥ 10 years)^62^ reference ranges. ^$^Travel by road estimated using GoogleMaps. ^†^Admission from acute medical or surgical ward. ^^^Central CRT > 2 seconds. ^+^GCS < 15 and/or AVPU < A. AVPU = Alert Voice Pain Unresponsiveness scale; CRT = capillary refill time; GCS = Glasgow Coma Scale.

| Clinical domain | Predictors | Ridge regression coefficient | p-value |
| --- | --- | --- | --- |
|  | Intercept | -1.8525 |  |
| Background | Age (months) | -0.0013 | 0.02 |
|  | Presence of comorbidity * | 0.2201 | 0.12 |
|  | Weight-for-age z-score (waz)<sup>#</sup> | -0.1108 | < 0.001 |
| Illness journey | Estimated travel time (minutes)<sup>§</sup> | 0.0013 | 0.01 |
|  | Intra-hospital transfer<sup>†</sup> | 0.4626 | < 0.001 |
| Cardiovascular | Heart rate (beats per minute) | -0.0002 | 0.89 |
|  | Heart rate (beats per minute) x Age (months) | 8.6514 x10<sup>-6</sup> | 0.14 |
|  | Prolonged capillary refill time<sup>^</sup> | 1.0244 | < 0.001 |
| Respiratory | Respiratory rate (beats per minute) | 0.0090 | 0.01 |
|  | Respiratory rate (beats per minute) x Age (months) | 4.8939 x10<sup>-5</sup> | 0.03 |
|  | Oxygen saturation (%) | -0.0253 | < 0.001 |
|  | Receipt of supplemental oxygen | 0.1808 | 0.10 |
| Neurological | Abnormal mental status<sup>+</sup> | 1.0313 | < 0.001 |

The model estimates the log odds of death during a PICU admission, using the sum of the intercept and the predictors multiplied by their coefficients, according to the following equation:
To support clinical decision-making, the output of the model (log odds) is converted into the probability that a PICU admission will result in death using the following transformation:

Discrimination of the new model (Figure 2a; AUC 0.84; 95% CI 0.80-0.88) was significantly better than all of the existing scores (DeLong test; p < 0.001). Calibration appeared best at lower predicted probabilities (Figure 2b), with the model underestimating risk for admissions with observed probabilities of death > 20-25%.

**Figure 2.**
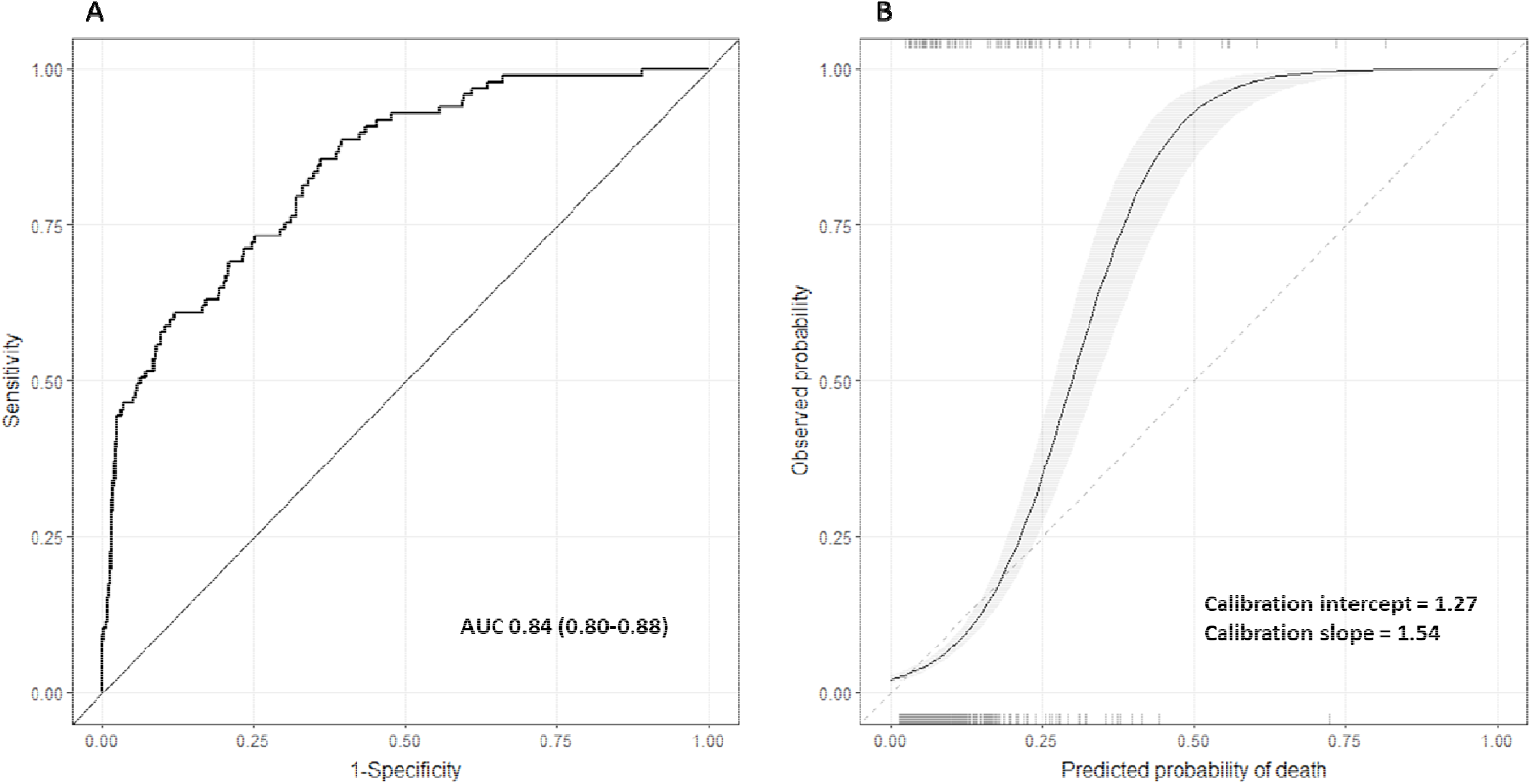
Discrimination and calibration of the new model. <u>Panel A</u>: discrimination of the new model. Perfect discrimination is indicated by an AUC of 1.0. <u>Panel B</u>: calibration of the new model. Dashed line indicates perfect calibration. Solid line indicates calibration of the model, with 95% confidence interval (grey ribbon). Rug plots indicate distribution of predicted risks for participants who did (top) and did not (bottom) meet the primary outcome. AUC = area under the receiver operating characteristic curve.

The ability of the model to triage PICU admissions into high- and low-acuity groups at cut-offs of 2.5%, 5%, 7.5%, 10%, and 15% is presented (Table 4). A cut-off of 10% reflects a triage strategy whereby all admissions with a predicted probability of death ≥ 10% are directed to a high-acuity area (where human and material resources are concentrated) and all other admissions are managed on the main unit. At this cut-off, admissions triaged to the high-acuity area would have a probability of death almost five times that of the general PICU population (PLR 5.75; 95% CI 4.57-7.23), whereas the probability amongst those triaged to the low-acuity area would be less than half that of the general PICU population (NLR 0.47; 95% CI 0.37-0.59), and almost a tenth of those triaged to the high-acuity area. At the 10% cut-off, approximately 13.0% of all admissions would be triaged to the high-acuity area, resulting in a ratio of 3:1 incorrect to correct (FP:TP) high-acuity triages.

**Table 4.** Ability of the model to triage PICU admissions. Performance of the model at five cut-offs (decision thresholds or threshold probabilities). A cut-off of 10% reflects a triage strategy whereby all admissions with a predicted probability of death ≥ 10% are directed to a high-acuity area and all other admissions managed on the main unit. A decrease in threshold probability (cut-off) is associated with an increase in the sensitivity of the triage strategy for identifying high-risk admissions, at the cost of a greater proportion of admissions being directed to the high-acuity area. FN = false negative (high-risk admission triaged to low-acuity area); FP = false positive (low-risk admission triaged to high-acuity area); NLR = negative likelihood ratio; PLR = positive likelihood ratio; TN = true negative (low-risk admission triaged to low-acuity area); TP = true positive (high-risk admission triaged to high-acuity area).

| Predicted probability of death | Sensitivity (95% CI) | Specificity (95% CI) | PLR (95% CI) | NLR (95% CI) | Per 1,000 admissions (~63 of which would die) |  |  |  | Percentage of admissions triaged as high-acuity | Ratio of incorrect to correct high-acuity triages |
| --- | --- | --- | --- | --- | --- | --- | --- | --- | --- | --- |
|  |  |  |  |  | TP | FP | TN | FN |  |  |
| 2.5% | 0.99<br>(0.97-1.00) | 0.16<br>(0.14-0.18) | 1.18<br>(1.14-1.21) | 0.07<br>(0.01-0.46) | 62 | 788 | 150 | 1 | 85.0% | 13:1 |
| 5% | 0.86<br>(0.79-0.93) | 0.63<br>(0.61-0.66) | 2.31<br>(2.08-2.57) | 0.23<br>(0.14-0.37) | 54 | 347 | 590 | 9 | 40.1% | 6:1 |
| 7.5% | 0.65<br>(0.56-0.74) | 0.80<br>(0.78-0.82) | 3.27<br>(2.73-3.91) | 0.44<br>(0.33-0.57) | 41 | 186 | 751 | 22 | 22.7% | 5:1 |
| 10% | 0.58<br>(0.48-0.68) | 0.90<br>(0.88-0.92) | 5.75<br>(4.57-7.23) | 0.47<br>(0.37-0.59) | 36 | 94 | 844 | 26 | 13.0% | 3:1 |
| 15% | 0.46<br>(0.37-0.56) | 0.96<br>(0.95-0.97) | 11.43<br>(8.22-15.88) | 0.56<br>(0.46-0.67) | 29 | 39 | 899 | 34 | 12.3% | 1:1 |

### Generalisability and applicability

There is great heterogeneity in the approach to critical care provision across different resource-constrained contexts, with the relative value of a TP and FP depending on the available human and material resources. Decision curve analyses accounting for these differing contexts indicate that using the model to support triage decisions could provide utility at cut-offs ≥ 7.5% (Figure 3), or simply put, in contexts where it might be desirable and feasible to manage up to a quarter (22.7%) of critical care admissions in a high-acuity area and tolerate up to 5:1 incorrect to correct (FP:TP) high-acuity triages.

**Figure 3.**
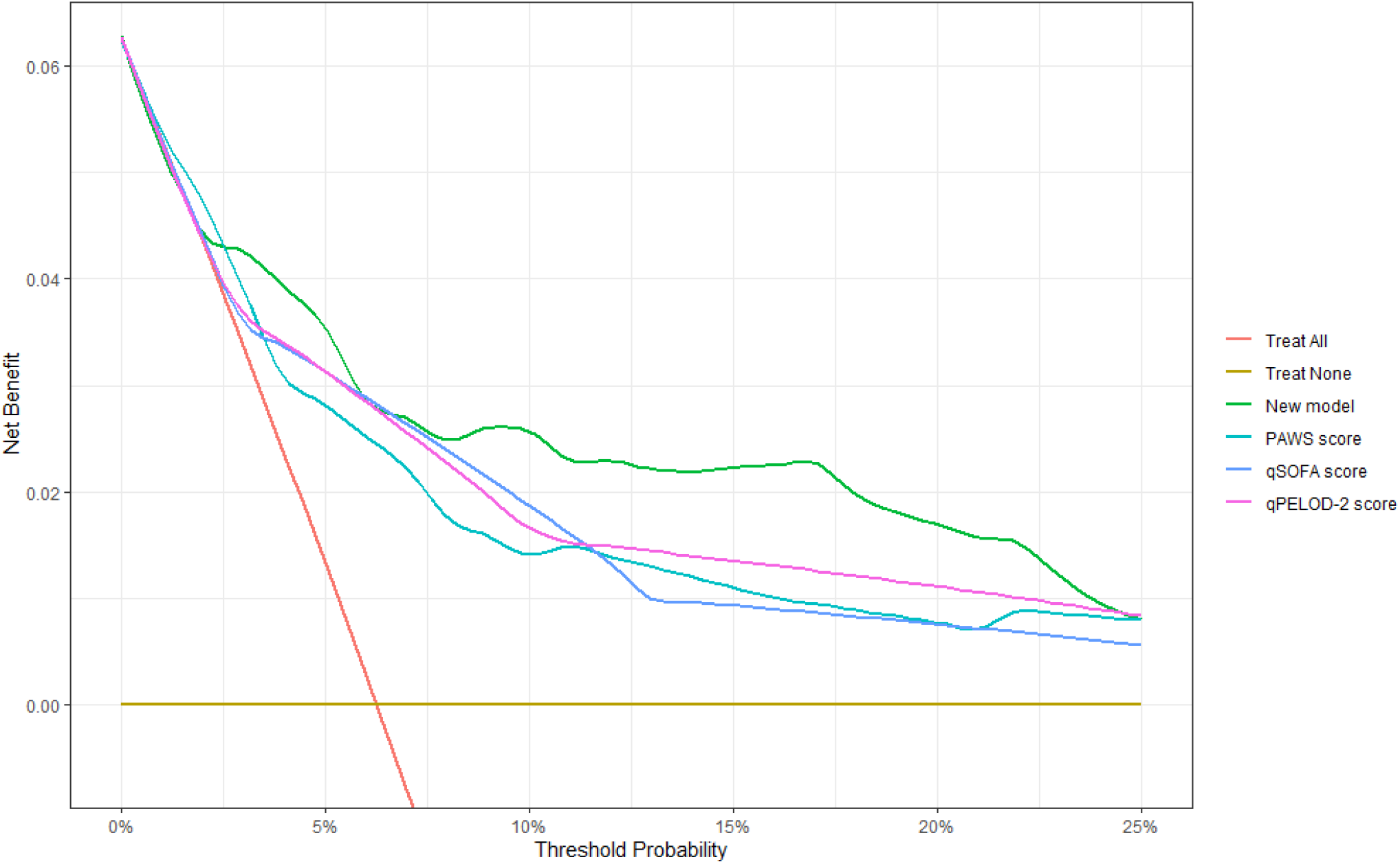
Clinical utility of the new model across a range of plausible decision thresholds. A cut-off (decision threshold or threshold probability) of 10% reflects a triage strategy whereby all admissions with a predicted probability of death ≥ 10% are directed to a high-acuity area and all other admissions managed on the main unit. The net benefit of the new model is compared to the three existing scores that demonstrated potential for stratifying admissions into low- and high-risk groups from the external validation. Above a cut-off of 7.5% using the new model to triage admissions appears to be the optimal strategy.

## Discussion

This study reports the ability of nine paediatric severity scores to risk stratify children on admission to an intensive care unit in Cambodia and compares their performance to that of a novel clinical prediction model derived specifically for locations where critical care resources are scarce. Whilst three scores (qPELOD-2, qSOFA, PAWS) appear to have moderate diagnostic utility, the new model proved superior and, if validated, could be a practical and flexible tool to support risk stratification of critically ill children in a variety of resource-constrained contexts.

With the exception of SIRS, which is known to be non-specific and perform poorly for risk assessment of acutely unwell children,^38, 39^ the other eight existing scores demonstrated comparable discrimination. However, discrimination is a poor indicator of clinical utility.^40^ Only three scores were associated with meaningful changes in the pre-test probability that a PICU admission might result in death, such that a single cut-off could be used to triage children into high- and low-risk groups. Whilst separate cut-offs could conceivably identify high- and low-risk admissions, in settings where resources are scarce it is unclear how a middle or ‘indeterminate’ group might be managed, and dividing admissions into multiple risk categories may not be practical on the ground.

Discrimination and classification of FEAST-PET, qSOFA, and qPELOD-2 were comparable to their original development studies,^38, 41, 42^ which may reflect similarities in the population (critically ill children), outcome (mortality), and for the FEAST-PET study, contextual factors (access to care, etc.). Performance of LqSOFA was inferior to the original development study,^43^ which is not unexpected as LqSOFA is known to perform best as a screening tool outside of PICU.^43, 44^ It is notable that discrimination and classification of PAWS, PEWS, and PEWS-RL were considerably worse in this study.^43, 45–47^ These scores are *diagnostic* scores (aiming to predict events occurring very shortly [< 24 hours] after the time of calculation) and it is therefore unsurprising that their ability to prognosticate more distal outcomes is sub-optimal.

The new model developed in this study considers the background of a child, their illness journey, and vital organ status to provide a contextualised assessment of critical illness and estimate the probability that a child will die prior to discharge, given the resources available in a typical Level II PICU located outside of a major urban centre in Southeast Asia. Discrimination of the model was considerably better than all existing severity scores evaluated. It provided good diagnostic value and was well calibrated over the threshold probabilities (cut-offs) of interest. In contexts where it might be feasible to resource a particular clinical area to manage up to a quarter of the highest risk admissions, if validated, the model could provide a readily implementable mechanism to identify children whom might benefit most from being cared for in such an area. Importantly, as the output of such a model is continuous (as opposed to discrete as is the case for points-based scores), the cut-off (threshold probability) for triage to the high-acuity area could be tailored to account for unit capacity, seasonal bed-pressures, hospital policy, and other dynamic contextual factors.

It is essential that risk stratification tools do not inadvertently concentrate all available resources on patients with untreatable and terminal illnesses.^1^ This is particularly important in contexts where resources are at a premium and prolonged hospitalisation can be associated with catastrophic expenditure for patients’ families. The PICU is often an environment where more resources are available to manage the end of life.^48^ Data-driven risk stratification can help reduce pressure on individual doctors and provide a framework for discussions related to dignified withdrawal of care.^8^

In addition to patient triage, accurate severity assessment tools, such as the one developed in this study, offer ancillary benefits. Severity-adjusted mortality rates can help standardise inter-unit comparisons and interpret impact of new interventions such as training programmes, therapeutics, or organisational changes.^49^ Risk scores and models can also improve transparency and consistency in the way policies are applied, and increase focus on care pathways to promote equitable and practical delivery of critical care.^50^ It is important to note that this risk prediction model is not intended to replace clinical assessment but rather to provide an additional data point to assist busy healthcare professionals plan and organise care for children who are critically ill.

This study provides one of few descriptions of paediatric critical care delivery in regions of LMICs where critical care demand and services are growing. Although preceding illness duration was reportedly short, children often had protracted journeys, consisting of multiple care encounters involving both the private and public health systems. The short average length of stay on PICU is striking and likely reflects the fact that in many resource-limited settings effective intensive care consists of providing simple, life-saving interventions for critically ill children with readily reversible conditions.^1, 4^ The ∼4% post-PICU discharge mortality rate is in keeping with other studies and is likely an underestimate due to considerable losses to follow-up.^51, 52^

Amongst this study’s strengths include it’s relatively unique setting in a PICU outside of a major urban tertiary centre in a country with high under-five mortality. Best-practice methods were followed for external validation of the existing scores and derivation of the new model, with particular care taken to prespecify and limit the number of candidate predictors and use penalised regression to avoid overfitting.^22^ Important contextual determinants of outcome absent in tools developed in high-income settings were included in the model and likely contributed to its promising performance.

The main limitation of this study is the lack of external validation of the new model. Although we took steps to avoid overfitting, assessment of the model’s performance in new data is required before it can be recommended for clinical use and to enable a fairer comparison with the pre-existing severity scores that were assessed in this study. A prospective validation study is underway.

Due to the retrospective nature of this study, travel time had to be estimated based on a child’s location of residence. This may not reflect actual travel time, which will be influenced by their mode of transport and interruptions to their journey. It was not possible to evaluate the performance of all longlisted scores. In particular, only two of the nine included scores were developed in LMICs and it is disappointing that seven LMIC-derived scores had to be excluded.^41, 47^

Almost 60% of the cohort were male children, which may reflect their predisposition to severe infections or gender biases in care seeking, although the latter is not known to be prevalent in Cambodia.^53, 54^ Nevertheless, the findings may be biased towards males. The use of routine records means that clinical parameters will not have been measured in a standardised manner. However, the existence of structured admission and vital signs proforma partially mitigate this issue and helped keep missingness low. Use of clinical records did ensure that the new model contains predictors feasible for collection under routine circumstances and will hopefully increase the likelihood of successful out-of-sample validation.

This study presents a new clinical prediction model for estimating the probability that a child admitted to a PICU in a resource-constrained context will not survive to discharge. The model contains predictors from multiple domains to ensure holistic assessment of critical illness. It outperformed nine existing paediatric severity scores and, if validated, offers a readily implementable and flexible mechanism to support risk stratification of critically ill children in resource-constrained contexts.

## Data Availability

De-identified, individual participant data from this study will be available to researchers whose proposed purpose of use is approved by the data access committees at the Angkor Hospital for Children and the Mahidol-Oxford Tropical Medicine Research Unit. Inquiries or requests for the data may be sent to.

## AUTHOR CONTRIBUTIONS

AC, SK, KP, AR, PT, NC, and CT conceptualised the study. AC shortlisted the existing severity scores. AC, SK, BS, CC, and HS collected the data. AC and SK reviewed the case report forms. AC, BS, CC, HS, and PV entered the data. PV conducted data entry checks. AR calculated the travel times. AC curated and cleaned the data. AC, SK, VM, NC, and CT selected the variables for the clinical prediction model. AC conducted the analysis under the supervision of LM, CK, and RPS. AC wrote the original draft of the manuscript. AC, SK, MV, BS, CC, HS, PV, KP, EH, AR, LM, CK, RPS, PT, NC, and CT reviewed, edited, and approved the manuscript. AC verified the underlying data.

## ACKNOWLEDGEMENTS

The authors are grateful to Soputhy Chansovannara and Phann Ysoun for their assistance with data collection, to Real Sophanith for her assistance with data entry, and to Thatsanun Ngernseng for setting up the study database.

