## Supplementary Appendix for "Derivation of a prognostic model for critically ill children in locations with limited resources"

### Derivation of a clinical prediction model to risk stratify paediatric intensive care admissions in locations with limited resources

#### SUPPLEMENTARY APPENDIX

**CONTENTS** **PAGE**


1. **TRIPOD checklist** 2
2. **Clinical proforma in routine use at the study site** 3
3. **Severity scores longlisted for external validation** 4
4. **Sensitivity analyses for primary outcome** 6
5. **Missing data patterns and results of sensitivity analyses** 7
6. **Study flowchart** 9
7. **Maps depicting location of residence of children admitted to PICU** 10
8. **UpSet plot illustrating clinical diagnoses amongst participants that died** 11
9. **Survival curves indicating time to meeting the primary and secondary outcomes** 12
10. **Calibration of existing severity scores** 13
11. **Sensitivity and specificity of existing severity scores** 14
12. **Relationship between continuous predictors and primary outcome** 15
13. **References** 16

#### Appendix 1. TRIPOD checklist.

| Section/Topic | | | Checklist Item | Page |
|---|---|---|---|---|
| **Title and abstract** | | | | |
| Title | 1 | D;V | Identify the study as developing and/or validating a multivariable prediction model, the target population, and the outcome to be predicted. | 1 |
| Abstract | 2 | D;V | Provide a summary of objectives, study design, setting, participants, sample size, predictors, outcome, statistical analysis, results, and conclusions. | 2 |
| **Introduction** | | | | |
| Background and objectives | 3a | D;V | Explain the medical context (including whether diagnostic or prognostic) and rationale for developing or validating the multivariable prediction model, including references to existing models. | 3 |
| | 3b | D;V | Specify the objectives, including whether the study describes the development or validation of the model or both. | 3 |
| **Methods** | | | | |
| Source of data | 4a | D;V | Describe the study design or source of data (e.g., randomized trial, cohort, or registry data), separately for the development and validation data sets, if applicable. | 4 |
| | 4b | D;V | Specify the key study dates, including start of accrual; end of accrual; and, if applicable, end of follow-up. | 4 |
| Participants | 5a | D;V | Specify key elements of the study setting (e.g., primary care, secondary care, general population) including number and location of centres. | 4 |
| | 5b | D;V | Describe eligibility criteria for participants. | 4 |
| | 5c | D;V | Give details of treatments received, if relevant. | 4 |
| Outcome | 6a | D;V | Clearly define the outcome that is predicted by the prediction model, including how and when assessed. | 6 |
| | 6b | D;V | Report any actions to blind assessment of the outcome to be predicted. | 4 |
| Predictors | 7a | D;V | Clearly define all predictors used in developing or validating the multivariable prediction model, including how and when they were measured. | 5-6 |
| | 7b | D;V | Report any actions to blind assessment of predictors for the outcome and other predictors. | 4 |
| Sample size | 8 | D;V | Explain how the study size was arrived at. | 6 |
| Missing data | 9 | D;V | Describe how missing data were handled (e.g., complete-case analysis, single imputation, multiple imputation) with details of any imputation method. | 6-7 |
| Statistical analysis methods | 10a | D | Describe how predictors were handled in the analyses. | 7 |
| | 10b | D | Specify type of model, all model-building procedures (including any predictor selection), and method for internal validation. | 7 |
| | 10c | V | For validation, describe how the predictions were calculated. | 7 |
| | 10d | D;V | Specify all measures used to assess model performance and, if relevant, to compare multiple models. | 7 |
| | 10e | V | Describe any model updating (e.g., recalibration) arising from the validation, if done. | N/A |
| Risk groups | 11 | D;V | Provide details on how risk groups were created, if done. | 7 |
| Development vs. validation | 12 | V | For validation, identify any differences from the development data in setting, eligibility criteria, outcome, and predictors. | Table 1 |
| **Results** | | | | |
| Participants | 13a | D;V | Describe the flow of participants through the study, including the number of participants with and without the outcome and, if applicable, a summary of the follow-up time. A diagram may be helpful. | 8 |
| | 13b | D;V | Describe the characteristics of the participants (basic demographics, clinical features, available predictors), including the number of participants with missing data for predictors and outcome. | 6-7, 8-9 |
| | 13c | V | For validation, show a comparison with the development data of the distribution of important variables (demographics, predictors and outcome). | N/A |
| Model development | 14a | D | Specify the number of participants and outcome events in each analysis. | 9 |
| | 14b | D | If done, report the unadjusted association between each candidate predictor and outcome. | N/A |
| Model specification | 15a | D | Present the full prediction model to allow predictions for individuals (i.e., all regression coefficients, and model intercept or baseline survival at a given time point). | Table 3 |
| | 15b | D | Explain how to the use the prediction model. | Table 3 |
| Model performance | 16 | D;V | Report performance measures (with CIs) for the prediction model. | 9-11 |
| Model-updating | 17 | V | If done, report the results from any model updating (i.e., model specification, model performance). | N/A |
| **Discussion** | | | | |
| Limitations | 18 | D;V | Discuss any limitations of the study (such as nonrepresentative sample, few events per predictor, missing data). | 13-14 |
| Interpretation | 19a | V | For validation, discuss the results with reference to performance in the development data, and any other validation data. | 12 |
| | 19b | D;V | Give an overall interpretation of the results, considering objectives, limitations, results from similar studies, and other relevant evidence. | 12-13 |
| Implications | 20 | D;V | Discuss the potential clinical use of the model and implications for future research. | 14 |
| **Other information** | | | | |
| Supplementary information | 21 | D;V | Provide information about the availability of supplementary resources, such as study protocol, Web calculator, and data sets. | 15 |
| Funding | 22 | D;V | Give the source of funding and the role of the funders for the present study. | 15 |

**Appendix 2. Clinical proforma in routine use at the study site.**

### Angkor Hospital for Children Admission Form

*Patient Label with Address*

*ICU 2022*

**Date:** ____/____/____ **Time**____________

**Doctor:** ______________________________________

**Chief Complaint**..........................................................................................................................................

**History** ..........................................................................................................................................................
.........................................................................................................................................................................
.........................................................................................................................................................................

**Past Medical History / Birth History** .............................................................................................................
.........................................................................................................................................................................
.........................................................................................................................................................................

**Current Medications**.........................................................................................................................................
.........................................................................................................................................................................

**Past Medications**..............................................................................................................................................
.........................................................................................................................................................................

**Allergies** .........................................................................................................................................................

**Yellow Card Reviewed:** ☐ Yes, ☐ No **Growth Chart Plotted:** ☐ Yes, ☐ No

**Immunizations:**

| (Birth) | (6weeks) | (10weeks) | (14weeks) | (6months) | (9months) | (18months) | |
|---|---|---|---|---|---|---|---|
| ☐ BCG<br>☐ $HepB_0$ | ☐ DPT- HepB $Hib_1$<br>☐ $PCV_1$<br>☐ $Polio_1$ | ☐ DPT- HepB $Hib_2$<br>☐ $PCV_2$<br>☐ $Polio_2$ | ☐ DPT- HepB $Hib_3$<br>☐ $PCV_3$<br>☐ $Polio_3$<br>☐ IPV | ☐ $MR_0$ | ☐ $MR_1$<br>☐ JE | ☐ $MR_2$ | ☐ Unknown<br>☐ Other:....<br>...............<br>................ |

**Development History**

**Gross Motor** head control-rolls-sits-crawls-walks-runs-stairs-jumps

**Fine Motor** eyes fix-reaches-transfers-unilateral reach-pincer-throws-dresses

**Speech/Hearing** alert to sound –coos-laughs-babbles-1 word-2 words-4 to 6 words-many words-asked questions

**Social** looks at face-social smile-recognizes parent-explores-uses spoons & cup-plays-continent urine and stool

Based on above is development appropriate for age? ☐ Yes, ☐ No.

**Further Development History**.........................................................................................................................

**Social History** ………………………………………………………………………………………………………………………………………………
……………………………………………………………………………………………………………………………………………………………
……………………………………………………………………………………………………………………………………………………………

**Family History**

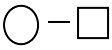

- female
- male

### Examination

Appearance ………………………………………………………………………………………………………………...

Temperature ……… ˚C HR.......... RR........... BP......./....... $O_2$ Sat......... RA or ....... L/mn $O_2$

Wt ……… kg Height ............ cm Wt/Ht Z-score: ...............SD

#### HEENT

Head Fontanelle..........................................................................Lymphadenopathy.....................
Eyes R…………………………………L……………………………..Fundus..................................
Ears R………………………….............L……………………………..
Nose
Throat
Other ..........................................................................................................................

#### Cardiovascular System

Pulse (circle) strong fair weak cannot detect
Heart Sounds ……………………………………………………………….
Capillary Refill Time ……………………………………………………………….
Other (JVP, precordium) .......................................................................................
.......................................................................................

#### Respiratory System

Auscultation ………..
Grunting/flaring ………..
Indrawing ………..

Other (percussion, tracheal position) ...........................................

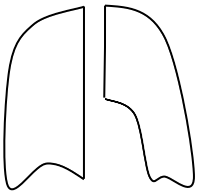

#### Gastro-Intestinal System

Inspection ………..
Palpation ………..
Organs ………..
Bowel Sounds ………..
Masses ………..

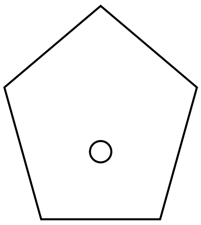

Other (genitalia, rectum, hernia, ascites) ............……………..............

**Nervous System**

Mental Status ........................ Kernig's ……………………

Neck Stiffness ……………….. GCS ………………………. (Use Chart Below)

Cranial Nerves ..................................................................................................................

| | 1 | 2 | 3 | 4 | 5 | 6 |
|---|---|---|---|---|---|---|
| EYES | Does not open | Opens in response to pain | Opens in response to voice | Opens spontaneously | X | X |
| VERBAL | Makes no sounds | Incomprehensible | Says inappropriate words | Confused and disoriented | Oriented and converses normally | X |
| MOTOR | No movements | Extension to pain | Abnormal flexion to pain | Flexion/Withdrawal to pain | Localizes painful stimuli | Obeys commands |

Peripheral Nerves **R** arm **L** arm **R** leg **L** leg

Tone…………………………………………………………………………...........

Power…………………………………………………………………………….

Reflexes………………………………………………………………………….

Sensation………………………………………………………………………..

Clonus ....................

Babinski ...................

Development ..........................................................................................................................

…………………………………………………………………… ….

**Skin & Extremities**

Rash ………………………………..

Edema ………………………………..

Wounds ………………………………..

Other ………………………………..

………………………………..

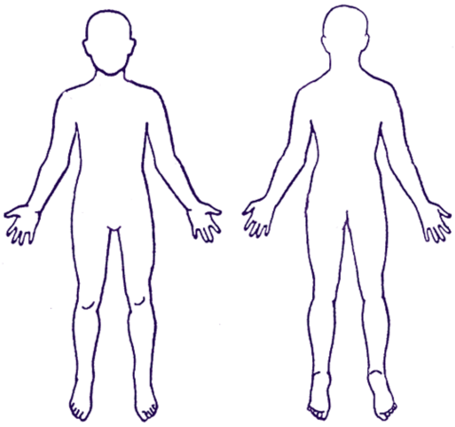

**Other**…………………………………………………………………………………………………..….

…………………………………………………………………………………………………………..….

…………………………………………………………………………………………………………..…

**Date:**........................................................**Bed Number:**............................**Name:**..................................................................................**Age**:............................................................

| Hours | | | 7 | 8 | 9 | 10 | 11 | 12 | 13 | 14 | 15 |
|---|---|---|---|---|---|---|---|---|---|---|---|
| **Pain / Sedation Assess** | **Temperature** | | | | | | | | | | |
| **NN** = No Need **PN** = Pain Need | **Patient Position** | | | | | | | | | | |
| **SN** = Sedation Need | **Heart Rate** | | | | | | | | | | |
| **GTT**= Continuous Drip | **NBP / ABP** | | | | | | | | | | |
| **Color** **Abd. Description** | **Color / MBP** | | | | | | | | | | |
| **PK** = Pink **S** = Soft | **CRT/ CVP** | | | | | | | | | | |
| **W** = Pale **T** = Tense | **Abd. Description / AG** | | | | | | | | | | |
| **C** = Cyanotic **D** = Distended | **Pain Scale Scores (1-10)** | | | | | | | | | | |
| **M** = Mottled | **Pain / Sedation** | | | | | | | | | | |
| **R** **Amount** **Tolerated** | **Set RR** | **Total RR** | | | | | | | | | |
| **E** **S** = Small **W** = Well | **SpO$_2$** % | **FIO$_2$** % | | | | | | | | | |
| **S** **M** = Medium **F** = Fair | **Mode** | | | | | | | | | | |
| **P** **L** = Large **P** = Poor | **P$_{PEAK}$ / P$_{MEAN}$** (CmH$_2$O) | | | | | | | | | | |
| **I** **1** = Thick **2** = Thin | **PEEP** (CmH$_2$O) | | | | | | | | | | |
| **R** | **I : E** ratio | | : | : | : | : | : | : | : | : | : |
| **A** **Description** | **I Time** / **T** $_{PL}$ | | | | | | | | | | |
| **T** **C** = Clear **Y**= Yellow | □TV □V$_{TE}$ □Insp TV (ml) | | | | | | | | | | |
| **O** **W**= White **B** = Brown | □Flow □V$_{ETOT}$ (LPM) | | | | | | | | | | |
| **R** **R** = Bloody **G** = Green | **Oral Care/Pt cir drain/O$_2$ check** | | | | | | | | | | |
| **Y** **Numeric pain Scale** | Suction | Amount | | | | | | | | | |
| **0** =No pain, **10**= worst pain | Tolerate | Describe | | | | | | | | | |
| P$_{PEAK}$: Peak Insp Pressure<br>P$_{MEAN}$: Mean Airway Pressure<br>T$_{PL}$: Plateau Time<br>V$_{ETOT}$: Exhaled Minute Volume<br>V$_{TE}$: Exhaled Tidal Volume | **NG Measurement Method**<br>1.Aspirate fluid from stomach<br>2.Listening by stetoscope<br>3.pH check ( Normal ≤5) | | | | | | | | | | |
| **GTTS:** | **mcg/mg \| kg \| hr/min** | | | | | | | | | | |
| **GTTS:** | **mcg/mg \| kg \| hr/min** | | | | | | | | | | |
| **GTTS:** | **mcg/mg \| kg \| hr/min** | | | | | | | | | | |
| **GTTS:** | **mcg/mg \| kg \| hr/min** | | | | | | | | | | |
| **F** **Method PO**=By mouth | **NG/OG/PO** | **Residual** | | | | | | | | | |
| **E** **Bt**=Bottle **BF**=Breastfed | **Amount In** | **Total** | | | | | | | | | |
| **E** **OG/NG**= Oral/Nasal Gastric | **Food type/Feed type** | | | | | | | | | | |
| **D** **C**: continuous **B**: Bolus | Post feeding position | | | | | | | | | | |
| | NG Measurement method | | | | | | | | | | |
| **IV Line Assessment** | **NG mark / Co-sign by TL/SN** | | | | | | | | | | |
| **S**= Soft **P** = Patent | 1 | 3 | | | | | | | | | |
| **L**= Locked **I** = Infiltrated | 2 | 4 | | | | | | | | | |
| **I** | Amount In | Total | | | | | | | | | |
| **N** | Amount In | Total | | | | | | | | | |
| **T** | Amount In | Total | | | | | | | | | |
| **A** | Amount In | Total | | | | | | | | | |
| **K** | Amount In | Total | | | | | | | | | |
| **E** | Amount In | Total | | | | | | | | | |
| **Blood Product1** | Amount In | Total | | | | | | | | | |
| **Blood Product 2** | Amount In | Total | | | | | | | | | |
| **O** **Stool** / **Right CT** | Amount Out | Total | | | | | | | | | |
| **U** Description / **Left CT** | Amount Out | Total | | | | | | | | | |
| **T** s = soft / **OG / NG** | Amount Out | Total | | | | | | | | | |
| **P** w = watery | Amount Out | Total | | | | | | | | | |
| **U** r = bloody / **Urine(ml)** | Amount Out | Total | | | | | | | | | |
| **T** l = loose / **Urine in cc/kg/h** | | | | | | | | | | | |
| m= mucus, bl= black / **Stool S M L** | Description | | | | | | | | | | |
| **Intake Output Balance** | | | | | | | | | | | |

### មន្ទីរពេទ្យកុមារអង្គរ

វិថីព្រះសង្ឃរាជ ទេព វង្ស និង ផ្លូវអុំឆាយ ភូមិមណ្ឌល ១ សង្កាត់ស្វាយដង្គំ ក្រុងសៀមរាប ប្រអប់សំបុត្រលេខ ៥០ ទូរស័ព្ទ: (៨៥៥) ៦៣ ៩៦៣ ៤០៩ ទូរសារ: (៨៥៥) ៦៣ ៧៦០ ៤៥២

Phreah Sangreach Tep Vong Road & Umchhay Street, Mondul 1, Sangkat Svay Dangkum P.O. Box 50, Siem Reap, Kingdom of Cambodia


**EMERGENCY / INTENSIVE CARE UNIT**
**Night time Pediatric Nursing Assessment**

Yesterday's weight……….Kg Bed Nº:………..
Today's Weight…………..Kg IBW… ………Kg
Days hospitalized………..Days in ICU…..........

Diagnosis: ____________________
History : ____________________
____________________
____________________
____________________
____________________
____________________
____________________

Ventilation: Yes☐ No☐
ETT size………Tape……….
Setting: Mode…….Rate…….
PIP/ PEEP……….. Ti………
FiO2…….. $V_T$…….$T_{pl}$……..
$V_{MAX}$ ______PS__________.
Plans …. ……………………
………………………………
………………………………
………………………………

#### ENVIRONMENTAL / SAFETY

☐ All alarms on, audible, and functioning ☐ Side rails up
☐ Pre-calculated drug sheet for patient weight
☐ Emergency equipments are at the bedside
☐ Hypothermic ☐ Hyperthermic Temp: ………..ºC

#### INTEGUMENTARY / MUSCULOSKELETAL — Normal ☐

☐Dry ☐Rash ☐Lesion ☐Breakdown ☐Petechiae ☐Thrush
☐Edema/swelling ☐Other problems:………………………………
Location: ………………………………………………………………

#### NEUROLOGICAL / FLACC PAIN SCALE — Normal ☐

**Anterior Fontanel** ☐ Bulging ☐ Sunken
**Movement of Extremities** ☐ Weak ☐ Absent ☐ Limit ROM
☐ Developmental Delay / CP ☐ Sedated ☐ Paralyzed
**GCS** Scale: ………../15
**Pupils reaction** ☐ Slow ☐ Fixed ☐ Dilated
☐ Unequal R:….….mm L:………mm
**Muscle tone** ☐ Hypertonic ☐ Hypotonic ☐ Quiet
☐ Flaccid ☐ Unresponsive
☐other…………………………………………………………………

| Category | Score 0 | Score 1 | Score 2 |
|---|---|---|---|
| **Face** | No particular expression or smile | Occasional grimace or frown, withdraw, disinterested | Frequent to constant frown, clenched jaw, quivering chin |
| **Legs** | Normal position or Relaxed | Uneasy Restless Tense | Kicking Or Legs drawn up |
| **Activity** | Lying quietly Normal position Moves easily | Squirming Shifting back/forth Tens | Arched Rigid Or Jerking |
| **Cry** | No Cry Awake or asleep | Moans or Whimpers Occasional complaint | Crying Steadily Screams or Sobs Frequent Complaint |
| **Consol ability** | Content Relaxed | Reassured by occasional touching, hugging or ' talking to' Distractible | Difficult to console or comfort |

#### COMFORT / SEDATION / IV SITE — Normal ☐

☐ Needs pain killer ☐ Needs sedation ☐ Needs paralysis
**Lines**: ☐ Erythema ☐ Skin damage ☐ Swelling
☐ Central ☐ Arterial ☐ IO
Location: ……………………Others………………………………

#### FLUIDS / NUTRITION — Normal ☐

☐ Abnormal
Patient is receiving total fluid. …………………………cc/kg/day
☐ Fluid restricted: ……………………………………………………
☐ Extra fluid:……………………………………………………………
☐ Abnormal Serum Electrolyte
Na: ……….. K: ……….. Ca: ……….. mmol/L ☐ Dr. Notified
☐ Fluid bolus / Blood products required:..…………………………..

#### OTHER ASSESSMENTS — Normal ☐

☐ Food support ☐ Abondoned child ☐ No parents visit
………………………………………………………………………

#### RESPIRATORY SYSTEM — Normal ☐

☐ Cough ☐ Dyspnea ☐ Slow Breathing
☐ Apnea ☐ Nasal Flaring ☐ Tachypnea
☐ Stridor ☐ Grunting ☐ Gasping
☐ Insufficient Respiratory Effort ☐ Thorax asymmetrical
**O2supply** ☐ Nasal Cannula ☐ NCPAP ……..$CmH_2O$
☐ Face Mask ☐ Face Mask with bag
☐ Flow: ……………….LPM
**Retraction** ☐ Suprastemal ☐ Supraclavicular
☐ Intercostal ☐ Subcostal
☐ Substernal
**Lungs' sound** ☐ Crackle ☐ Left ☐ Right
☐ Wheezing ☐ Left ☐ Right
☐ Decreased ☐ Left ☐ Right
☐ Other sound: …………………………..........
**Ventilator**: ETT Size: …………mm Depth: …………...cm
ET Suction size: ..…….Fr Depth: …………...cm
Mode: ……………………PIP/PEEP: …………….cmH2O
Rate: …………. FiO2: …………% TV: ……………….
IT: ………..sec Other settings: ………………..………
**Chest tube**: Size: ……..Fr ☐ Right ☐ Left ☐ Suction
☐ Gravity ☐ Chest fluid color: ……………………
**Others**: …………………………………………………………..……

#### CARDIOVASCULAR SYSTEM — Normal ☐

☐ Tachycardia ☐ Bradycardia ☐ Murmur ☐ CRT>2sec
☐ Hypertensive ☐ Hypotensive
☐ Active precordium ☐ BP not detectable
☐ Cool extremities Last Hct:………..% On …………………..
☐ Sweating ☐ JVP
**Color**: ☐ Mottled ☐ Pallor ☐ Jaundice
**Pulse**: ☐ Weak ☐ Unequal ☐ Not palpable
**Cyanosis**: ☐ Peripheral ☐ Central
**Others**: …………. ……………………………………………………..

#### GASTROINTESTINAL / GENITOURINARY — Normal ☐

☐ Vomiting ☐ Constipation ☐ AG…...cm
☐ Poor appetite ☐ Colostomy ☐ NPO
☐ Imperforate Anus
**Abdomen** ☐ Pain ☐ Firm ☐ Distended
☐ Hepatomegaly ☐ Spleen palpable
**Bowel sound** ☐ Hypoactive ☐ Hyperactive ☐ Absent
**Stool** ☐ Watery ☐ Mucusy ☐ Bloody
☐ Loose ☐ Black
**Feeding Tube** ☐ Oral ☐ Nasal size: …..Fr, Day……...
**Urine** ☐ Urine<1ml/kg/h ☐ Urine>5ml/kg/h ☐ Anuria
☐ Cloudy ☐ Dark Yellow ☐ Hematuria
**Foley catheter** Size: ……………Fr, Day…………
**Others**: ………………………………………………………………..

#### ASSESSMENT COMPLETED BY

**Initial** :.. ………..**Signature**:………………… ….**Time**:………………

**Appendix 3. Severity scores excluded at longlisting.** Severity scores identified from two recent systematic reviews and PubMed search. Scores were excluded if they contained advanced diagnostic tests unlikely to be available in resource-constrained contexts, included variables that were not relevant for the intended setting of use, the information required to calculate the score/model was not provided in the original manuscript, or the required variables were not available in the routine clinical records at the study site (and no suitable proxy variable could be identified). APTT = activated partial thromboplastin clotting time; ARI = acute respiratory infection; BUN = blood urea nitrogen; GE = gastroenteritis; HIV = human immunodeficiency virus; LDH = lactate dehydrogenase; IPSCC = international pediatric sepsis consensus conference; MUAC = mid-upper arm circumference; $paCO_2$ = partial pressure of carbon dioxide in arterial blood; $paO_2$ = partial pressure of oxygen in arterial blood; PCT = procalcitonin; PICU = paediatric intensive care unit; PT = prothrombin time; RA = room air; $SpO_2$ = oxygen saturation.

| NAME OF SCORE | ADVANCED DIAGNOSTIC TEST REQUIRED | INAPPROPRIATE FOR SETTING AND/OR POPULATION | DATA NOT AVAILABLE | REASONS FOR EXCLUSION |
|---|---|---|---|---|
| **AQUAMAT** | Y | N | N | BUN and base deficit required |
| **BITWE MODEL** | Y | N | Y | MUAC, infectious diagnosis (ARI, GE, malaria, bacteraemia, other) required |
| **BITWE SCORE** | Y | N | Y | MUAC, infectious diagnosis (ARI, GE, malaria, bacteraemia, other) required |
| **DRAMAIX** | Y | N | Y | Albumin, transthyretin, oedema, and MUAC required |
| **ELSHOUT** | N | Y | N | Sore throat, palpable lymphadenopathy not suitable for PICU population |
| **ERDMAN** | Y | N | N | Host biomarker tests required |
| **FEAST-PETaL** | Y | N | N | BUN, pH, and lactate required |
| **ITAT** | N | Y | N | $SpO_2$ on RA not relevant for PICU population |
| **KWIZERA 1** | N | N | N | Information not available for construction of the score/model |
| **KWIZERA 2** | N | N | N | Information not available for construction of the score/model |
| **KWIZERA 3** | N | N | N | Information not available for construction of the score/model |
| **KWIZERA 4** | N | N | N | Information not available for construction of the score/model |
| **KWIZERA 5** | N | N | N | Information not available for construction of the score/model |
| **LIN NOMOGRAM** | Y | N | N | Blood culture, albumin, and LDH required |
| **LODS** | N | N | Y | Deep breathing and prostration required |
| **LOWLAAVAR 1** | N | Y | N | HIV not relevant (low endemicity) |
| **LOWLAAVAR 2** | N | Y | Y | HIV not relevant (low endemicity), MUAC required |

| | | | | |
|---|---|---|---|---|
| **LOWLAAVAR 3** | N | N | Y | MUAC required - and as this is a model cannot substitute a proxy variable |
| **MPIMBAZA** | N | N | Y | Prostration, jaundice, deep breathing, and meningitic signs required |
| **mPRIO** | Y | Y | Y | $SpO_2$ on RA, PCT, organ dysfunction as per IPSCC definition |
| **mRISC** | N | Y | Y | Malaria not relevant (low endemicity), dehydration, prostration, night sweats, and historical loss of consciousness required |
| **PCIS** | Y | N | Y | BUN, creatinine, $K^+$, $Na^+$, pH, $paO_2$, and gastrointestinal bleeding required |
| **PEDIA-e** | N | N | Y | Prostration, jaundice, and kwashiorkor required |
| **PEDIA-i** | N | N | Y | Deep breathing, prostration, and jaundice required |
| **PEDIA-l** | N | N | Y | Prostration and kwashiorkor required |
| **PELOD-2** | Y | N | Y | Lactate, creatinine, $paO_2$, $paCO_2$, and pupillary reaction required |
| **PERCH** | N | Y | Y | $SpO_2$ on RA not relevant, deep breathing, cough, and grunting required |
| **PEWS BCH** | N | N | Y | Skin colour, frequency of nebulisation prior to assessment, nurse concern, and family concern required |
| **PIM III** | Y | Y | Y | Base excess, PaO2, and pupillary reactions required, along with many other high-income country contextual variables |
| **PIRO** | Y | Y | Y | $paO_2$, BUN, transaminases, PT, blood culture, $SpO_2$ on RA not relevant for PICU population, signs of liver failure |
| **pMODS** | Y | N | N | Lactate, bilirubin, $paO_2$, fibrinogen, and BUN required |
| **PRISM III** | Y | N | Y | $pCO_2$, $paO_2$, pH, acidosis, total $CO_2$, $K^+$, BUN, creatinine, PT/APTT, and pupillary reflexes required |
| **pSOFA** | Y | N | N | Bilirubin and creatinine required |
| **qSOFA-L** | Y | N | N | Lactate required |
| **RISC** | N | Y | Y | $SpO_2$ on RA not relevant for PICU population and HIV not relevant (low endemicity), prostration, and wheezing required |
| **RISC-Malawi** | N | Y | Y | $SpO_2$ on RA not relevant for PICU population, MUAC, and wheezing required |
| **SCOTT** | N | Y | Y | Arrival via emergency medical services, indwelling central line, and hospitalised within last year required |
| **SICK** | N | Y | N | $SpO_2$ on RA not relevant for PICU population |
| **TORPS** | N | Y | N | $SpO_2$ on RA not relevant for PICU population |
| **YOS** | N | N | Y | Quality of cry, reaction to parent stimulation, state variation, colour, and response to social overtures required |

**Appendix 4. Sensitivity analyses for primary outcome.** Top table: comparison between the primary analysis (n = 1,550; outcome events = 97) and a sensitivity analysis (n = 1529; outcome events = 76) where admissions which met the primary outcome but in which the death was judged to have been related to a second illness acquired during the PICU stay (n = 9) and admissions which were discharged to die at home (n = 12) were excluded. Bottom table: comparison between the primary analysis and a sensitivity analysis in which laboratory parameters were restricted to those available between two hours prior and four hours after PICU admission.[1] Results are presented for the two scores which included laboratory parameters: FEAST-PET (haemoglobin) and SIRS (white cell count). In the sensitivity analysis, with the more restrictive criteria for inclusion of laboratory parameters, missingness (addressed by median imputation grouped by outcome status as for the primary analysis) increased to 30.2% for FEAST-PET and 31.0% for SIRS.

| MODEL / SCORE | AUC (95% CI) | |
|---|---|---|
| | PRIMARY ANALYSIS | SENSITIVITY ANALYSIS |
| NEW MODEL | 0.84 (0.80-0.88) | 0.83 (0.78-0.88) |
| FEAST-PET | 0.72 (0.66-0.78) | 0.74 (0.67-0.80) |
| LqSOFA | 0.76 (0.71-0.81) | 0.79 (0.74-0.84) |
| PAWS | 0.76 (0.71-0.81) | 0.77 (0.72-0.82) |
| PEWS | 0.71 (0.65-0.76) | 0.72 (0.66-0.79) |
| PEWS-IRISH | 0.74 (0.69-0.79) | 0.74 (0.69-0.80) |
| PEWS-RL | 0.72 (0.67-0.77) | 0.72 (0.67-0.78) |
| qPELOD-2 | 0.75 (0.70-0.80) | 0.76 (0.70-0.81) |
| qSOFA | 0.74 (0.69-0.79) | 0.74 (0.69-0.80) |
| SIRS | 0.59 (0.53-0.65) | 0.62 (0.55-0.68) |

| MODEL / SCORE | AUC (95% CI) | |
|---|---|---|
| | PRIMARY ANALYSIS | SENSITIVITY ANALYSIS |
| FEAST-PET | 0.72 (0.66-0.78) | 0.72 (0.66-0.78) |
| SIRS | 0.59 (0.53-0.65) | 0.58 (0.52-0.63) |

**Appendix 5. Missing data patterns and results of sensitivity analyses.** Top figure: missingness pattern for existing severity scores evaluated in external validation. Bottom figure: missingness pattern for candidate predictors included in new clinical predicition model. Table: results of sensitivity analyses conducted using different approaches for handling missing data. For best case imputation, missing values amongst admissions that met the primary outcome were assigned the most extreme values in the dataset, whilst missing values amongst admissions that did not meet the primary outcome were assigned a normal value (e.g. median heart rate, 100% oxygen saturation, no supplemental oxygen, etc.) The opposite approach was taken for worst case imputation, with missing values amongst admissions that met the primary outcome being assigned a normal value and missing values amongst admissions that did not meet the primary outcome being assigned an extreme value.

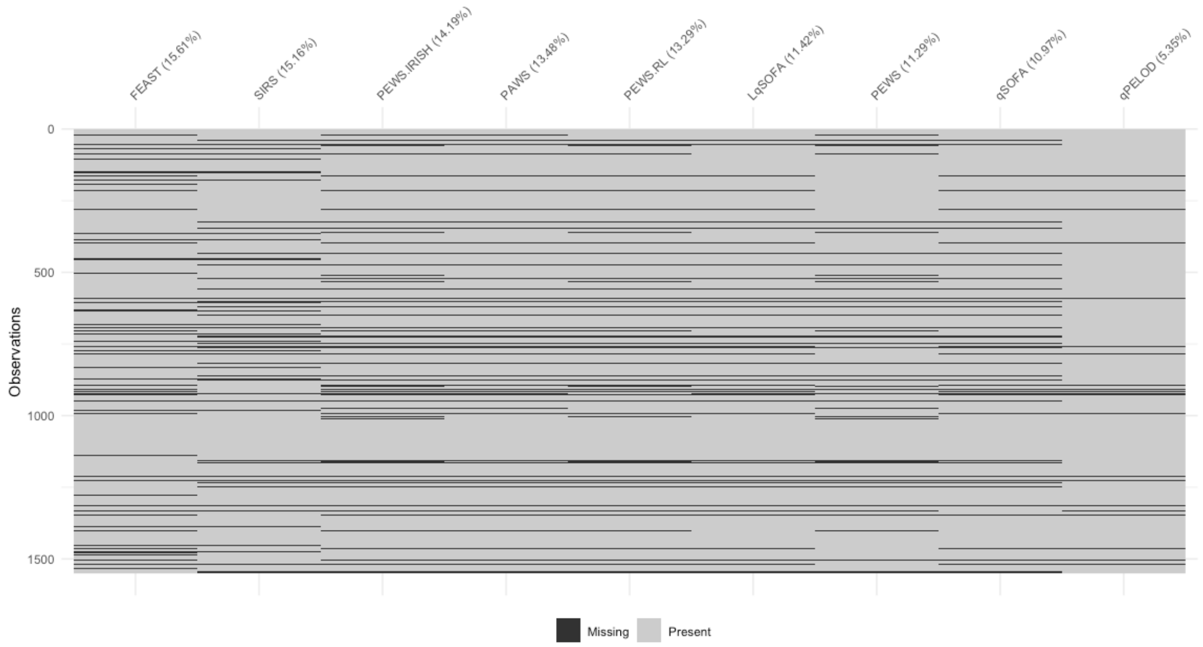


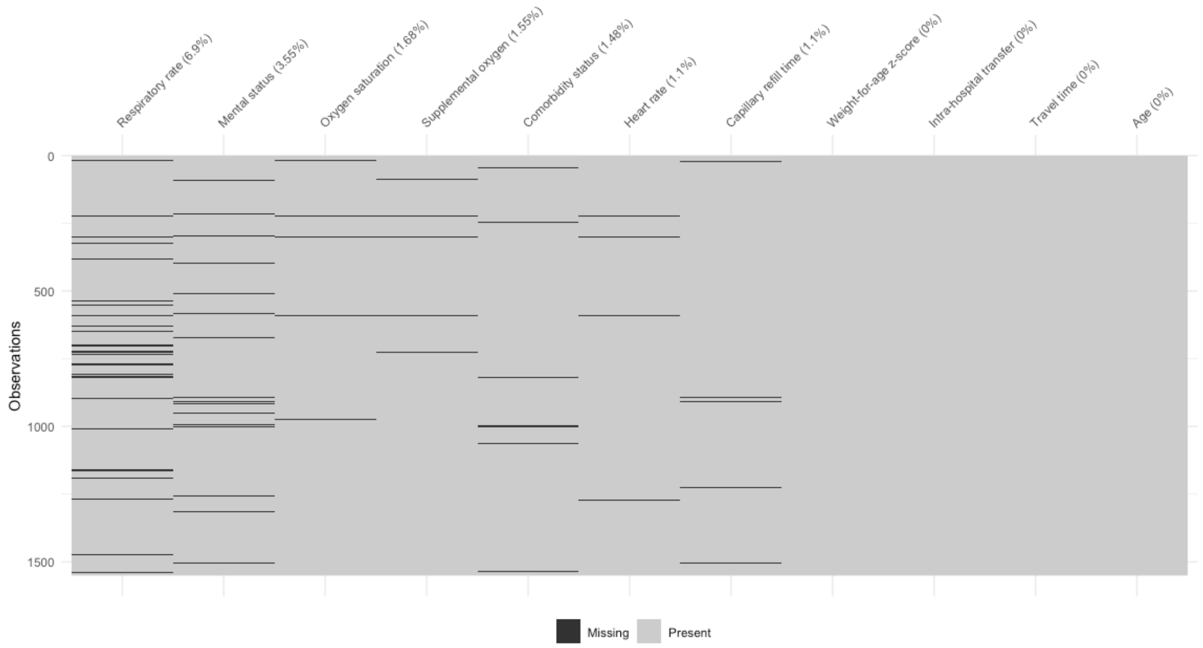

| MODEL / SCORE | AUC (95% CI) | | | |
|---|---|---|---|---|
| | COMPLETE CASE | MEDIAN IMPUTATION | BEST CASE | WORST CASE |
| **NEW MODEL** | 0.83 (0.79-0.88) | 0.84 (0.80-0.88) | 0.86 (0.83-0.90) | 0.78 (0.74-0.83) |
| **FEAST-PET** | 0.67 (0.60-0.74) | 0.72 (0.66-0.78) | 0.74 (0.68-0.80) | 0.67 (0.61-0.73) |
| **LqSOFA** | 0.75 (0.69-0.80) | 0.76 (0.71-0.81) | 0.78 (0.73-0.83) | 0.71 (0.66-0.76) |
| **PAWS** | 0.73 (0.67-0.78) | 0.76 (0.71-0.81) | 0.78 (0.73-0.83) | 0.70 (0.65-0.75) |
| **PEWS** | 0.67 (0.61-0.73) | 0.71 (0.65-0.76) | 0.73 (0.67-0.78) | 0.67 (0.62-0.73) |
| **PEWS-IRISH** | 0.69 (0.63-0.75) | 0.74 (0.69-0.79) | 0.76 (0.71-0.81) | 0.69 (0.64-0.74) |
| **PEWS-RL** | 0.68 (0.62-0.74) | 0.72 (0.67-0.77) | 0.73 (0.68-0.78) | 0.66 (0.61-0.71) |
| **qPELOD-2** | 0.73 (0.68-0.79) | 0.75 (0.70-0.80) | 0.75 (0.70-0.80) | 0.70 (0.64-0.75) |
| **qSOFA** | 0.72 (0.66-0.77) | 0.74 (0.69-0.79) | 0.74 (0.69-0.79) | 0.70 (0.64-0.75) |
| **SIRS** | 0.62 (0.55-0.68) | 0.59 (0.53-0.65) | 0.63 (0.57-0.68) | 0.56 (0.50-0.62) |

**Appendix 6. Study flowchart.**

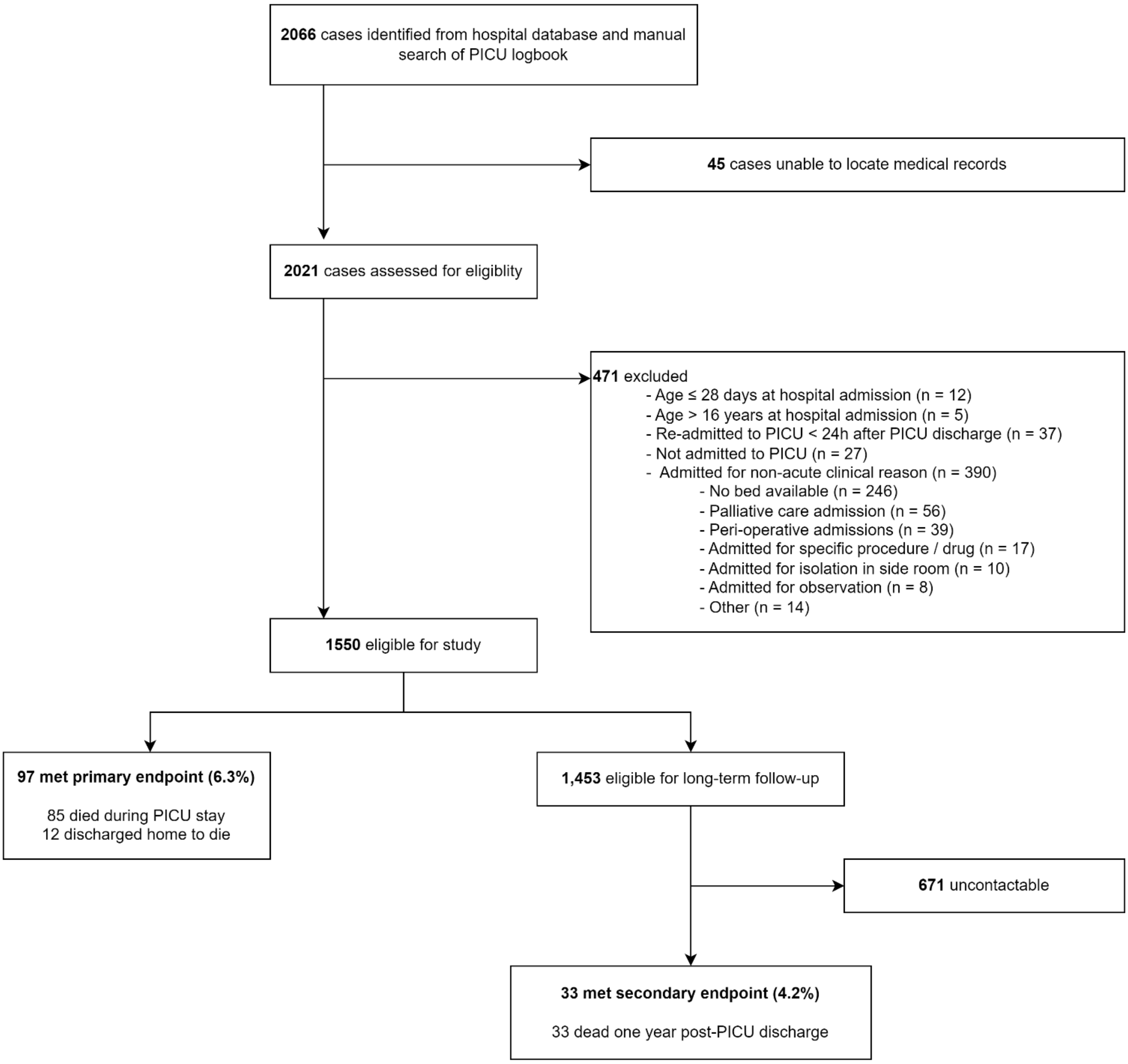

**Appendix 7. Maps depicting locations of residence for children admitted to PICU.** Left panel: distribution of admissions across Cambodian provinces. Right panel: distribution of admissions across communes in Siem Reap province. The study site is indicated by the red dot.

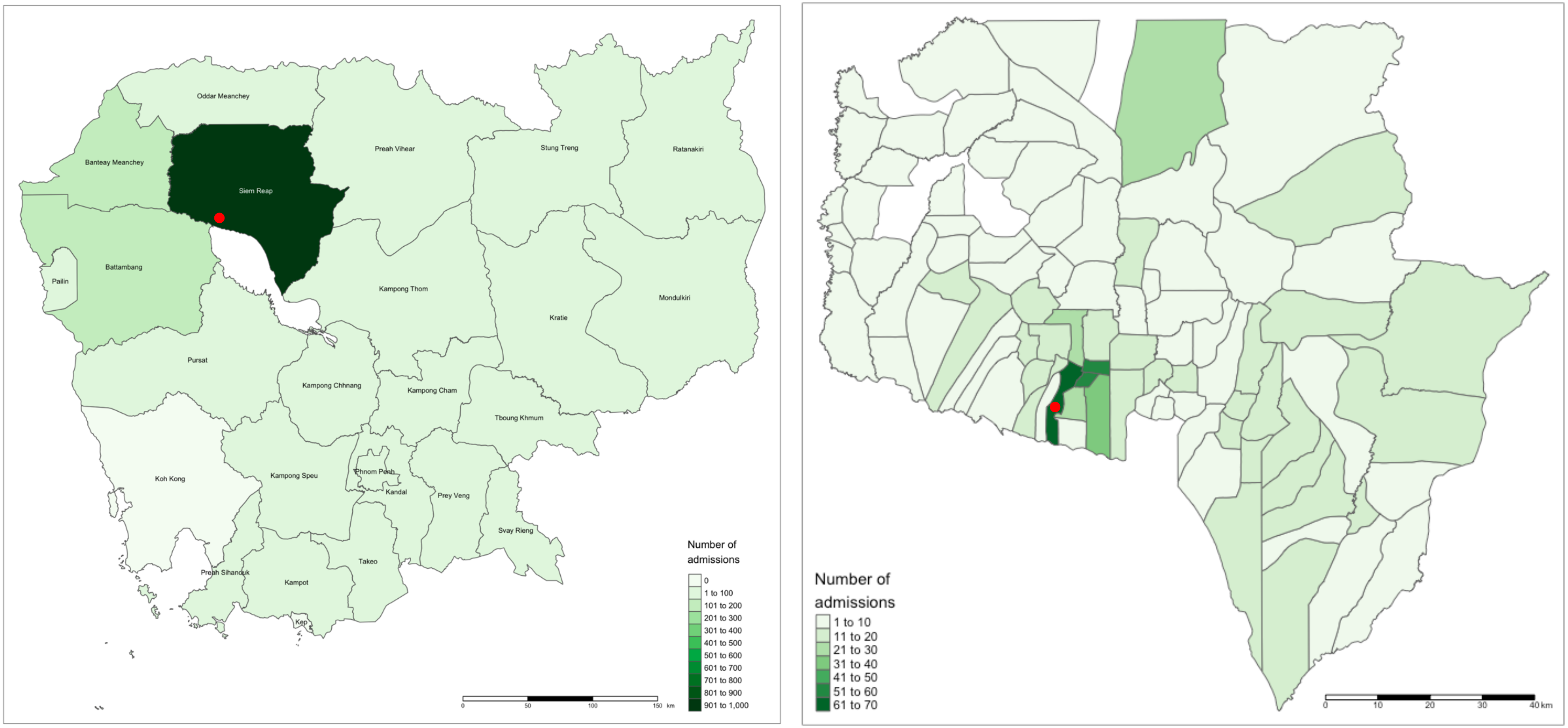

**Appendix 8. UpSet plot illustrating clinical diagnoses amongst participants that died.** Sets contain children with a particular diagnosis. Intersections contain children with a particular combination of diagnoses.

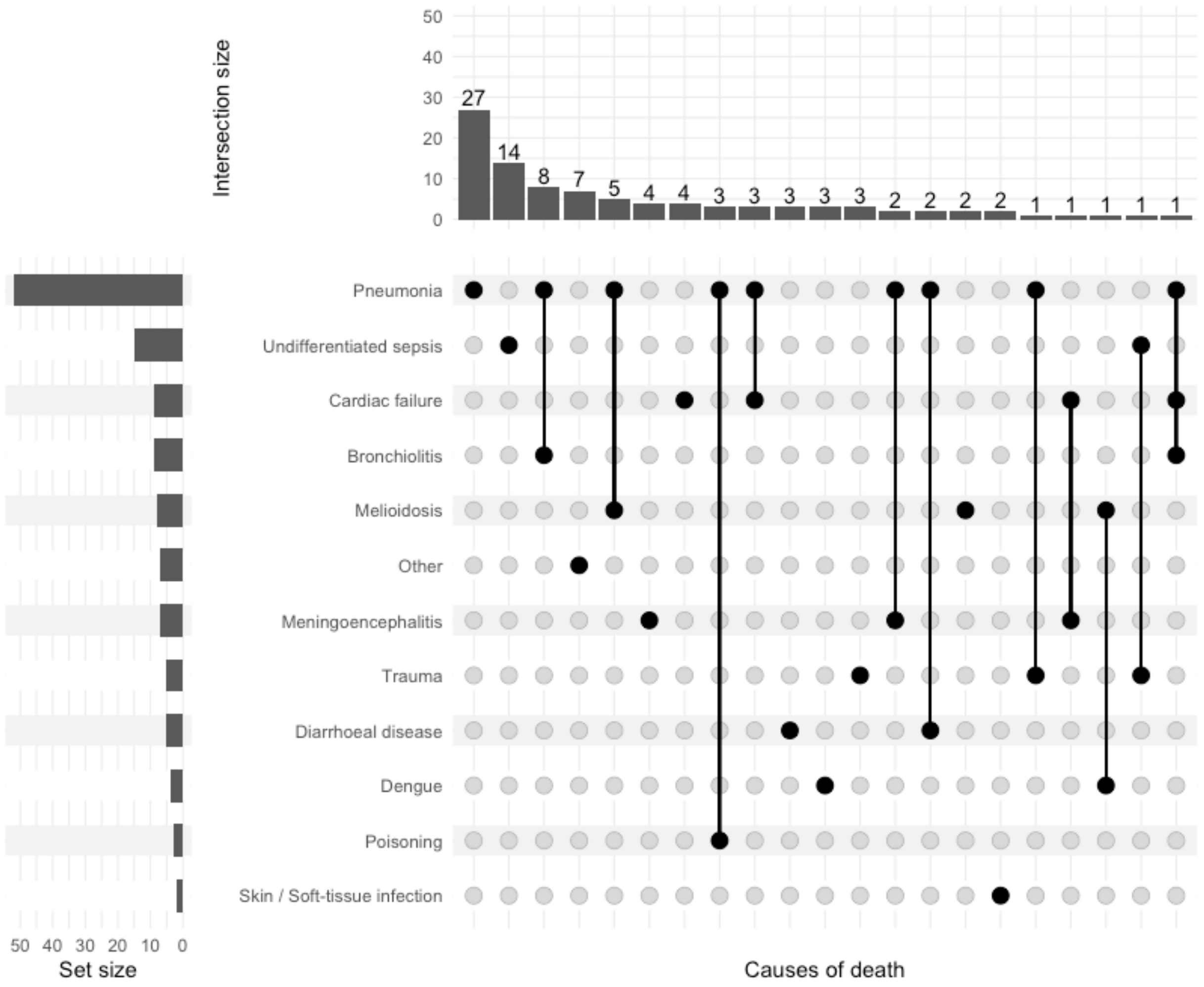

**Appendix 9. Time to meeting the primary and secondary outcomes.** Upper panel: days to death after PICU admission depicted on a histogram (left) and survival curve (right); n = 1,550. Lower panel: months to death after PICU discharge depicted on a histogram (left) and survival curve (right); n = 782. Grey ribbons indicate 95% confidence intervals.

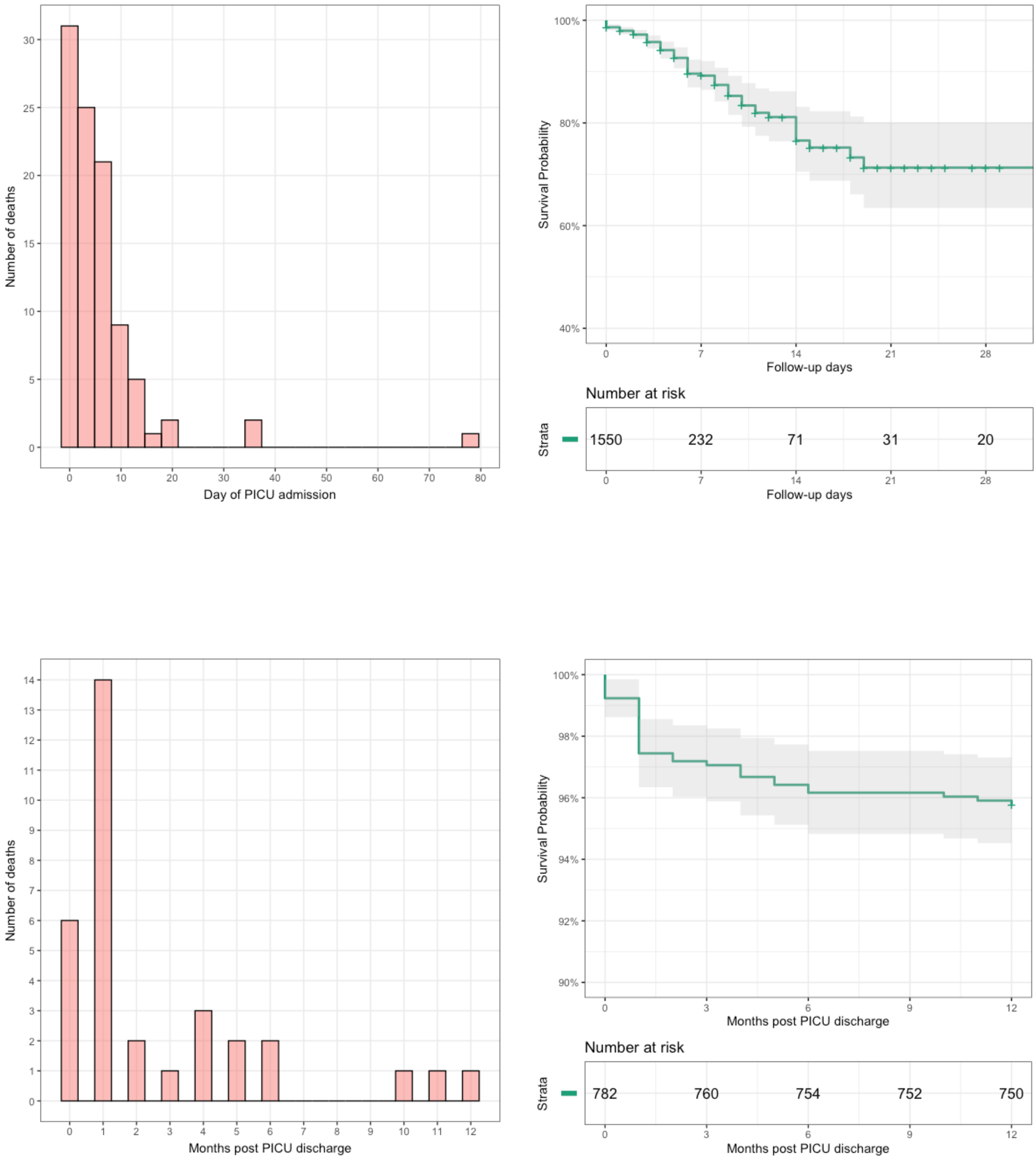

**Appendix 10. Calibration of existing severity scores.** Proportion of admissions at each level of each score that died during their PICU stay. Error bars indicate Wilson 95% confidence intervals.

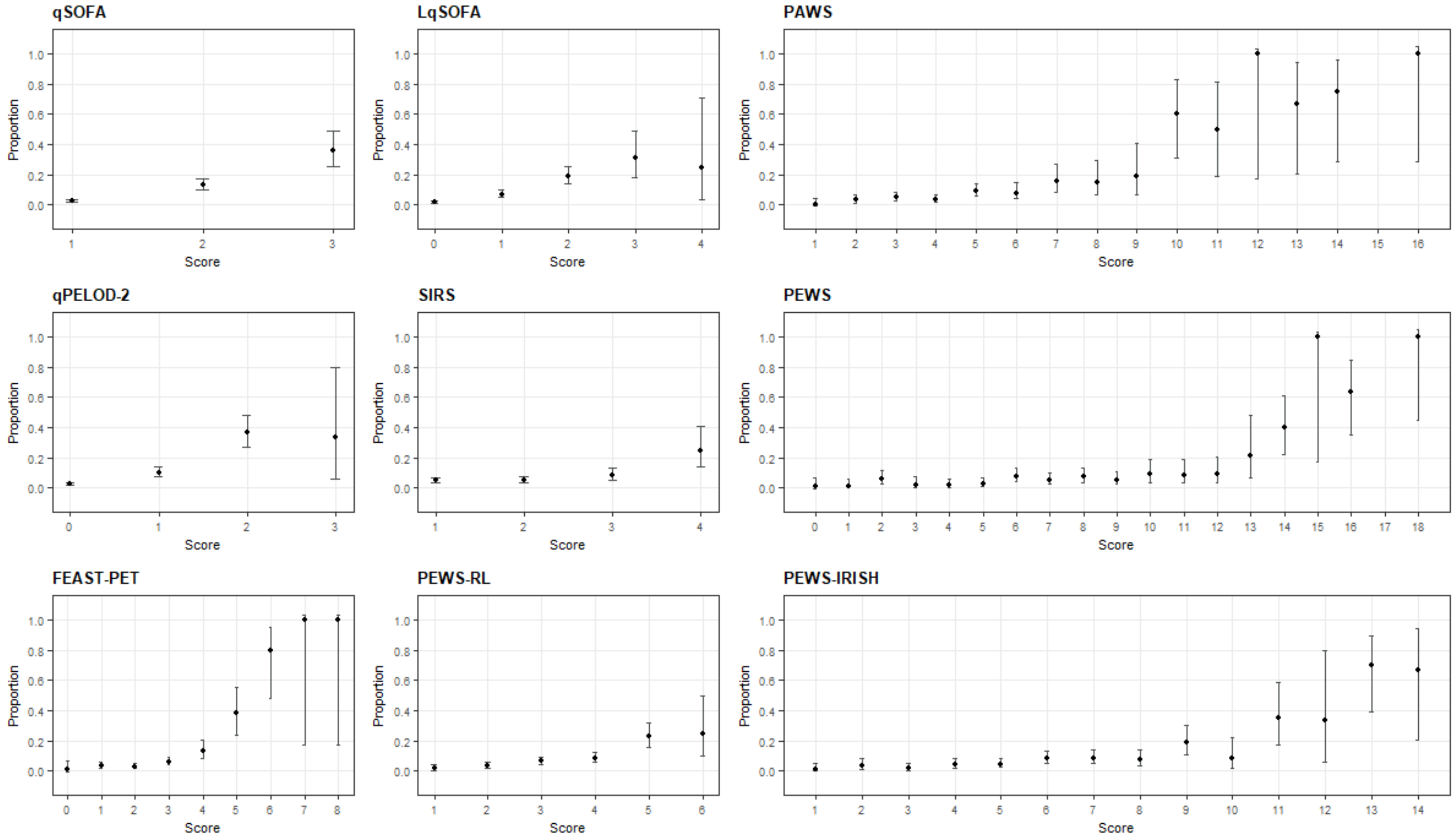

**Appendix 11. Sensitivity and specificity of existing severity scores.** Change in sensitivity (red line) and specificity (blue line) at increasing cut-offs of the severity scores. Grey shaded ribbons indicate 95% confidence intervals.

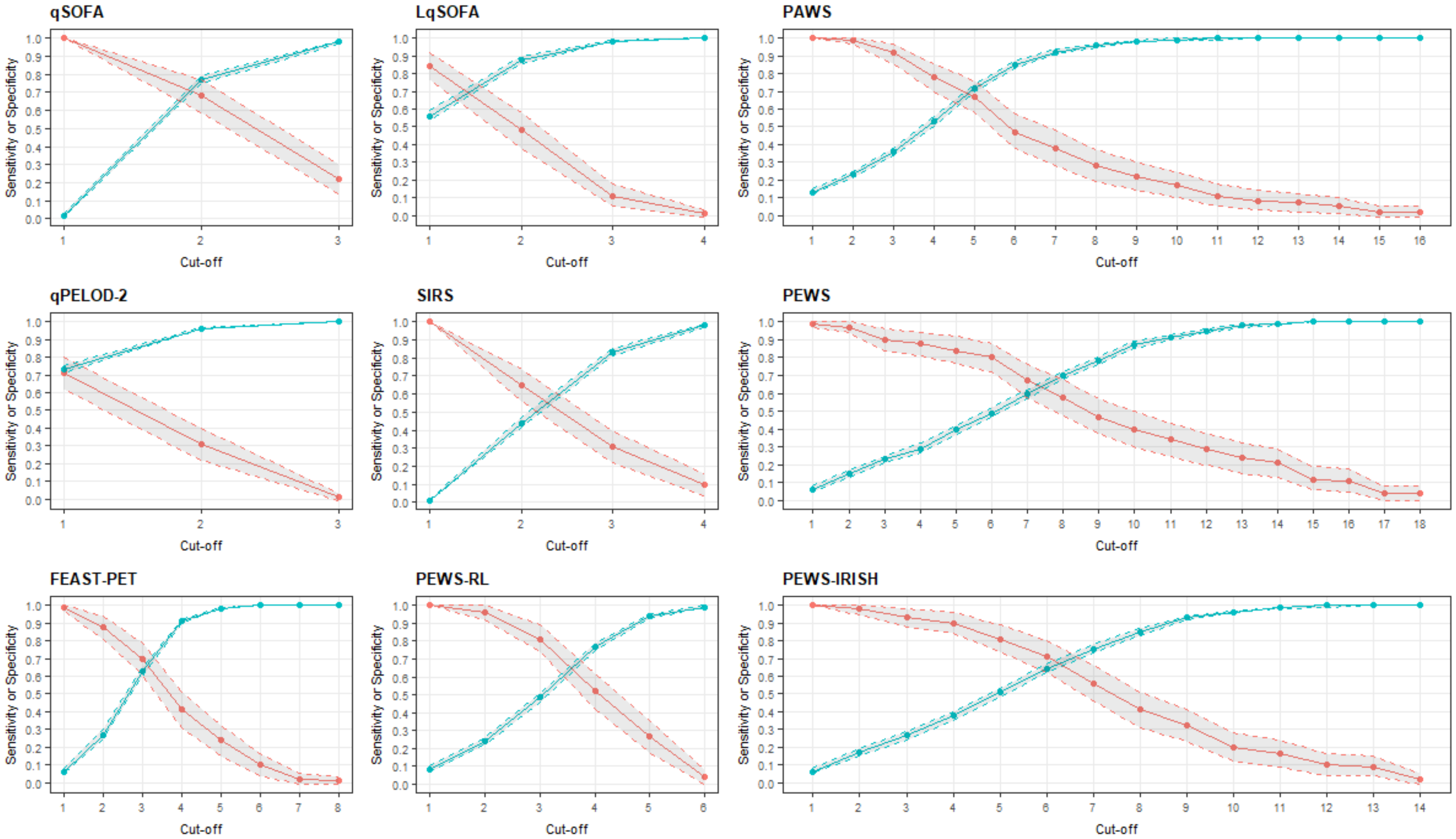

**Appendix 12. Relationship between continuous candidate predictors and the primary outcome.** Loess smoothed curves to explore the relationship between continuous candidate predictors and the probability of death during PICU admission to determine if transformations might be required for the modelling. Grey ribbons indicate 95% confidence intervals.

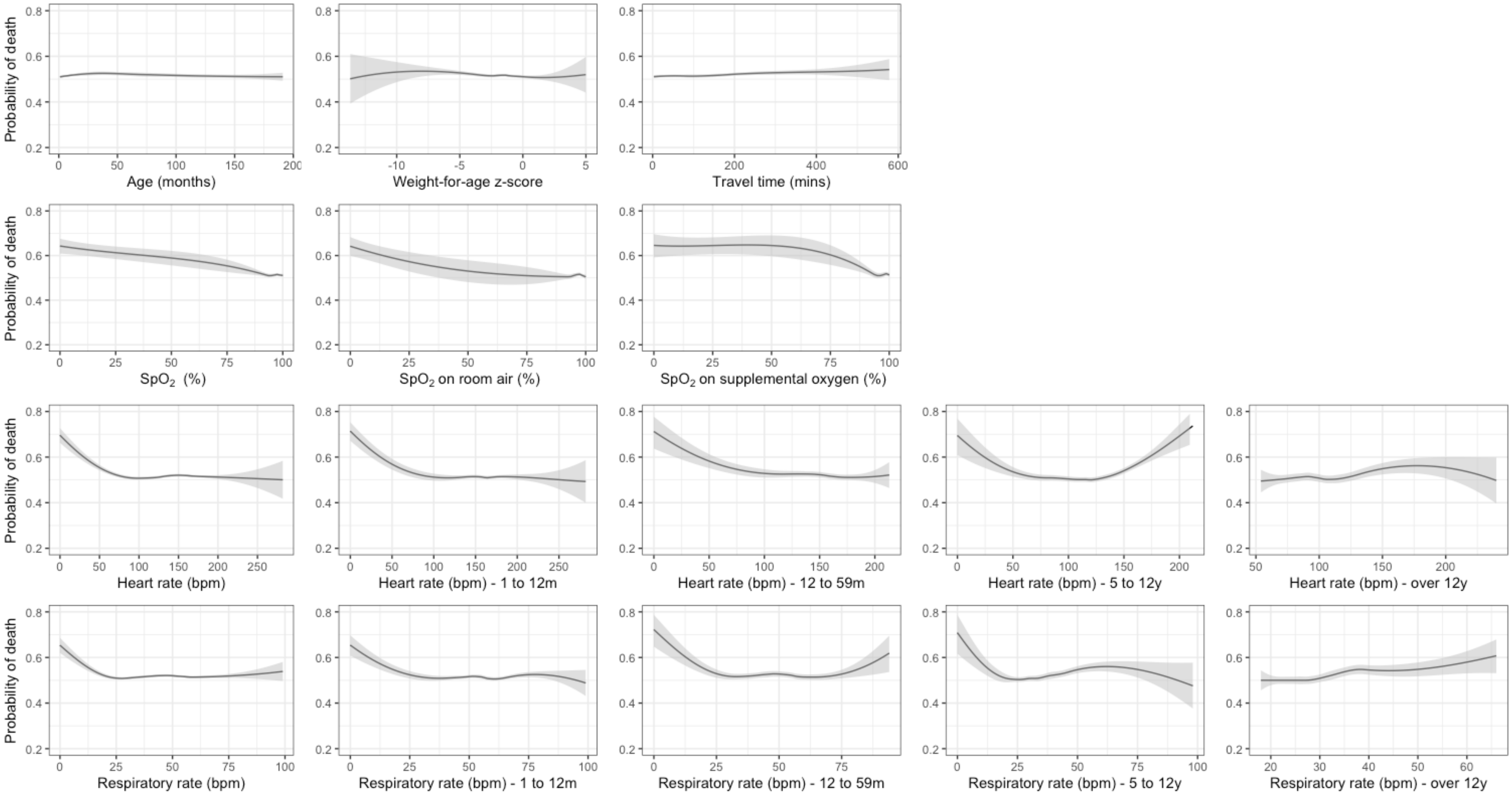
